# Screen Time and Puberty Timing: Findings from the Adolescent Brain Cognitive Development Study

**DOI:** 10.64898/2026.01.26.26344850

**Authors:** Luise Bläschke, Franka Edith Weisner, Anke Hinney, Triinu Peters, Börge Schmidt, Raphael Hirtz, Lars Dinkelbach

## Abstract

**Objective:** To examine whether screen time predicts interindividual variability regarding pubertal development across adolescence.

**Study design:** This longitudinal cohort study included 10786 participants (47.9% female) from the Adolescent Brain Cognitive Development (ABCD) study. First, associations were examined between average daily screen time (hours/day, parent-reported Screen Time Survey) at baseline (mean age = 9.91 ± 0.63 years) and pubertal timing, derived from Pubertal Development Scale (PDS) scores through 4-year follow-up (mean age = 14.08 ± 0.68 years) and standardized by age and sex. Second, associations were examined between screen time groups (very low: 0–1.29 h/day; low: 1.29–2.07 h/day; moderate: 2.07–2.86 h/day; high: 2.86–4.0 h/day; very high: 4.00–12.43 h/day) and age at mid-puberty, defined as the age at first parent report of Pubertal Development Scale (PDS) category at least 3.

**Results:** In linear mixed models adjusting for age, sex, race/ethnicity, socioeconomic status, BMI, and physical activity, higher log-transformed screen time at baseline was associated with more advanced pubertal timing at 1-, 2- and 3- year follow-ups, with the strongest effect observed at year 2 (standardized ß=0.07 [95%-CI, 0.05 to 0.10]). The associations were more pronounced in girls. The group of participants with very high screen time reached mid-puberty 2.47 months earlier [adjusted effect size, 95%-CI, -3.38 to -1.56) than participants with very low screen time.

**Conclusion:** These findings suggest that screen time in late childhood is linked with earlier pubertal development and highlight its relevance for parental guidance on preadolescents’ screen media use.

## Introduction

Screen devices such as smartphones, tablets, televisions, and computers play an increasingly prominent role in the daily lives of children and adolescents, whether for leisure, social interaction, or education. Multiple adverse associations of screen use with youth health have been reported, including higher BMI [1, 2], increased cardiometabolic and cardiovascular risk [3], impaired sleep [4], and depressive symptoms [5].

Alongside increased screen use, there is a trend toward earlier onset of puberty: In girls, the age at thelarche decreased world-wide by approximately three months per decade between 1977 and 2013 [6]. A similar secular trend toward earlier puberty has been observed in a study of Swedish boys [7]. In particular, during the COVID 19 pandemic, several countries reported a consistent rise in pathologically accelerated pubertal development in girls, including higher incidence rates of central precocious puberty [8-12]. Among the proposed mechanisms, such as reduced physical activity and increased body weight during lockdowns, increased exposure to screen light and digital devices has been suggested as a contributing factor [10, 13, 14]. This hypothesis is supported by both experimental work in rodents and observational studies in humans. In rats, exposure to blue light at wavelengths similar to those emitted by LED screens has been shown to induce earlier pubertal onset in females [15] and males [16], an effect which has been replicated after exposure to computer or mobile phone screens compared with exposure to artificial warm fluorescent lighting [17]. In line with this, a recent meta analysis of ten observational studies, including only two longitudinal studies, found that higher screen time treated as a continuous measure was associated with earlier pubertal development, whereas analyses using categorical thresholds (≤2 h/day versus ≥2 h/day) and sex-stratified subgroups did not yield significant associations [18]. Given these heterogeneous findings, the inconsistent control of confounders, and the limited number of longitudinal designs, the authors emphasized the need for more rigorous longitudinal research before firm conclusions can be drawn.

Here, we address these gaps using longitudinal data from the ABCD Study, a population-based US cohort that examines physical and psychosocial development in children and adolescents, to assess whether screen time exposure in late childhood (ages 9–11 years) is (1) associated with pubertal timing at follow-up assessments through age 14, (2) related to the age at which mid-pubertal stages are reached, and (3) clarify sex specific patterns of the association between screen time and puberty timing in sex-stratified analyses.

## Methods

### Sample Description and Exclusion Criteria

The ABCD Study is a prospective cohort study of 11868 participants recruited across 21 sites in the United States. For the present analyses, data from baseline (age 9.91±0.63 years) through the 4-year follow-up (age 14.08±0.68 years) were analyzed.

Participants were excluded if they had missing or implausible baseline demographic data (age, race/ethnicity, body mass index standard deviation score (BMI SDS)), missing or implausible exposure data (screen time at baseline), missing outcome data (pubertal development information at baseline and/or at all assessments), or an unclassifiable sex assigned at birth (mismatch between PDS-reported sex and sex assigned at birth or an intersex designation, Supplementary Figure S1). The final study sample comprised 10786 participants (47.89% female) (Table 1, Supplementary Table S1, Supplementary Table S2). The number of participants with complete follow up data at each time point was as follows: year 1, N=9686 (89.80% of the initial sample; age 10.92±0.64 years; 47.68% female); year 2, N=9363 (86.81%; age 12.02±0.67 years; 47.35% female); year 3, N=8694 (80.60%; age 12.91±0.65 years; 46.81% female); and year 4, N=4047 (37.52%; age 14.08±0.68 years; 46.65% female).

**Table 1.** Characteristics of Participants by Screen Time Group.

| Screen Time Group | Very Low<br>N=1695 | Low<br>N=1695 | Moderate<br>N=1695 | High<br>N=1695 | Very High<br>N=1695 |
| --- | --- | --- | --- | --- | --- |
| Range of Screen Time,<br>Hours per Day | (0.0 – 1.29) | (1.29 – 2.07) | (2.07 – 2.86) | (2.86 – 4.00) | (4.00 – 12.43) |
| **Demographic Information** |  |  |  |  |  |
| Female, % | 66.73% | 59.35% | 57.94% | 53.77% | 52.27% |
| Age at Baseline ,<br>mean (SD) | 9.95 (0.61) | 9.95 (0.62) | 10.00 (0.61) | 9.99 (0.62) | 10.01 (0.63) |
| **Race/Ethnicity, N (%)** |  |  |  |  |  |
| White | 1090 (64.31) | 1018 (60.06) | 939 (55.40) | 760 (44.84) | 607 (35.81) |
| Black | 112 (6.61) | 157 (9.26) | 189 (11.15) | 294 (17.35) | 472 (27.85) |
| Hispanic | 254 (14.99) | 308 (18.17) | 368 (21.71) | 419 (24.72) | 407 (24.01) |
| Asian | 71 (4.19) | 43 (2.54) | 31 (1.83) | 22 (1.30) | 23 (1.36) |
| Other | 168 (9.91) | 169 (9.97) | 168 (9.91) | 200 (11.80) | 186 (10.97) |
| **Weight Category<sup>a</sup>, N (%)** |  |  |  |  |  |
| Underweight | 72 (4.25) | 64 (3.78) | 46 (2.71) | 56 (3.30) | 42 (2.48) |
| Normal Weight | 1280 (75.52) | 1170 (69.03) | 1070 (63.13) | 978 (57.7) | 907 (53.51) |
| Overweight | 191 (11.27) | 233 (13.75) | 281 (16.58) | 314 (18.5) | 303 (17.88) |
| Obese | 152 (8.97) | 228 (13.45) | 298 (17.58) | 347 (20.5) | 443 (26.14) |
| **Screen Time, Hours per Day** |  |  |  |  |  |
| At Baseline, mean<br>(SD) | 0.88 (0.35) | 1.71 (0.21) | 2.46 (0.20) | 3.35 (0.35) | 6.11 (2.35) |
| At 2 Year Follow-Up,<br>mean (SD) | 2.25 (1.87) | 2.99 (2.06) | 3.63 (2.29) | 4.39 (2.61) | 5.46 (3.23) |
| Baseline Social Media<br>Use, mean (SD) | 0.05 (0.27) | 0.09 (0.35) | 0.10 (0.34) | 0.16 (0.48) | 0.20 (0.56) |
| **Total Family Income, N<sup>b</sup> (%)** |  |  |  |  |  |
| < \$5000 | 51 (3.01) | 59 (3.48) | 44 (2.60) | 72 (4.2) | 104 (6.14) |
| \$5,000 - \$11,999 | 34 (2.01) | 54 (3.19) | 54 (3.19) | 69 (4.1) | 124 (7.32) |
| \$12,000 - \$15,999 | 28 (1.65) | 26 (1.53) | 34 (2.01) | 44 (2.6) | 71 (4.19) |
| \$16,000 - \$24,999 | 45 (2.65) | 66 (3.89) | 98 (5.78) | 81 (4.8) | 136 (8.02) |
| \$25,000 - \$34,999 | 67 (3.95) | 95 (5.60) | 85 (5.01) | 128 (7.6) | 141 (8.32) |
| \$35,000 - \$49,999 | 80 (4.72) | 133 (7.85) | 136 (8.02) | 181 (10.7) | 230 (13.57) |
| \$50,000 - \$74,999 | 188 (11.09) | 208 (12.27) | 237 (13.98) | 255 (15.0) | 248 (14.63) |
| \$75,000 - \$99,999 | 204 (12.04) | 233 (13.75) | 263 (15.52) | 286 (16.9) | 223 (13.16) |
| \$100,000 - \$199,999 | 666 (39.29) | 581 (34.28) | 551 (32.51) | 457 (27.0) | 336 (19.82) |
| > \$200,000 | 332 (19.59) | 240 (14.16) | 193 (11.39) | 122 (7.2) | 82 (4.84) |

To enhance interpretability across varying levels of screen use, participants were categorized into five equally sized groups based on their daily screen time: very low (0.0–1.29 h/day), low (1.29–2.07 h/day), moderate (2.07–2.86 h/day), high (2.86–4.00 h/day), and very high (4.00–12.43 h/day). As the focus of this differentiation was the effect of each screen time group on age at mid-puberty, this classification was conducted in participants with available information on age at midpuberty (N=8475, 78.57% of the study sample).

### Screen Time

Participants’ screen time at baseline was assessed using the Parent Screen Time Survey. Parents were instructed to report their child’s daily use of screen devices such as computers, cellphones, tablets, or other electronic devices on typical weekdays and weekend days, excluding time spent on school-related activities. Average daily screen time was calculated as a weighted mean, assuming 5 weekdays and 2 weekend days. Screen time values exceeding 24 hours per day were considered implausible and treated as missing. In line with previous studies examining the impact of screen time in the ABCD study, extreme values above the 99th percentile (12.43 h/day, N=106 (<1.00%)) were winsorized to the 99th percentile ([19], Supplementary Figure S2). To enhance interpretability across varying levels of screen use, participants were categorized into five equally sized groups based on their daily screen time: very low (0.0–1.29 h/day), low (1.29–2.07 h/day), moderate (2.07–2.86 h/day), high (2.86–4.0 h/day), and very high (4.0–12.43 h/day).

### Puberty Timing

Pubertal development was evaluated using the parent-reported Pubertal Development Scale (PDS). For primary analyses, the parental ratings were used as these show higher agreement with clinical assessments than self-reports in this age group [20, 21]. The PDS includes five items on physical pubertal characteristics, covering sex-neutral (growth spurt, body hair, skin changes) and sex-specific characteristics (facial hair growth and voice deepening in males; breast development and menarche in females). At each follow-up (years 1–4), relative pubertal timing was estimated by regressing mean PDS scores on chronological age separately by sex [22]; the resulting standardized residuals indexed pubertal timing, with positive values indicating earlier and negative values indicating later development compared with same-aged, same sex peers (Supplementary Figure S3, S4).

### Age at Mid-Puberty (PDS Category 3)

The PDS allows for reliable differentiation between prepubertal (stage 1–2), mid-pubertal (stage 3), and postpubertal (stage 4–5) categories [23]. The age at the first parent report of at least mid-puberty (PDS category ≥3), here referred to as “age at mid-puberty”, was derived from the five assessments spanning baseline to the four-year follow-up. Due to missing PDS data at later follow-ups, this information was available in only N=8475 participants (57.92% female; 78.57% of the study sample) who reached mid-puberty during the study.

### Statistical Analyses

#### Analysis of Continuous Screen Time and Puberty Timing

Based on visual inspection (Supplementary Figures S5 and S6) and model fit assessed using the Akaike Information Criterion (AIC, Supplementary Table S3), a natural log transformation of screen time was applied, as it was most appropriate for modeling its association with pubertal timing. After evaluating residual normality (Supplementary Figure S7), linearity (Supplementary Figures S5, S6), and absence of multicollinearity (Supplementary Table S4), crude and adjusted linear mixed models were run to estimate the effect of baseline screen time on pubertal timing at follow up. In crude models, the effect of log-transformed screen time on pubertal timing was estimated with random intercepts for site and for families nested within sites. The adjusted models additionally included fixed effects for demographic factors (sex, age at baseline, race/ethnicity), socioeconomic factors (parental education, family income, area deprivation index), and physical factors at baseline (physical activity and BMI SDS). These covariates were selected based on a directed acyclic graph (DAG) to determine the minimal sufficient adjustment set to estimate the total causal effect of screen time on puberty timing (Supplementary Figure S8). Details on the operationalization of covariates are provided in the Supplementary Methods.

To estimate the effects of screen time groups (very low to very high, with the very low group as reference) on age at mid-puberty, crude and adjusted linear mixed models using the same covariate set as in the continuous screen time analyses were run.

Missing data for education (N=10 [<0.1% of study sample]), income (N=820 [7.60%], area deprivation index (N=790 [7.32%]), and physical activity (N=1653 [15.33%]) were imputed using multiple imputation (Supplementary Methods Covariates and Missingness, Supplementary Table S5-S10, Supplementary Figures S9, S10).

### Sensitivity Analyses

To test the robustness of our findings with respect to exposure definitions, variable transformations, and handling of missing data, we conducted the following sensitivity analyses: (1) using youth reported instead of parent reported screen time, (2) using untransformed rather than log-transformed screen time, and (3) excluding participants with missing data on parental education, income, area deprivation index, or physical activity rather than imputing these values (Supplementary Methods Sensitivity Analyses, Supplementary Figure S11).

### Additional Analyses

To assess the contribution of individual covariates, we ran sequential models for log-transformed screen time, adding one covariate at a time. Adjusted models were also stratified by sex to examine potential sex-specific associations with pubertal timing. Additional models evaluated the effect of log-transformed screen time on puberty tempo, defined as the change in parent-reported PDS score from baseline to year 2 relative to the corresponding change in age. Given the increasing relevance of social media use, we also examined associations between youth-reported social media use and parent-reported pubertal timing. Because information on other sources of screen time was not collected, potential differences across media types could not be evaluated.

## Results

Adjusted linear mixed effects models revealed that higher baseline log-transformed screen time was associated with earlier pubertal timing at subsequent follow-ups (Figure 1, Figure 2). The association between standardized, log-transformed baseline screen time and standardized pubertal timing (both sexes combined) was strongest at the 2-year follow-up, where a one standard deviation increase in log-transformed screen time was associated with a 0.07 standard deviation earlier pubertal timing (β=0.07 [95%-CI, 0.05 to 0.10]), Supplementary Table S11). The association was similar at the 1-year follow-up (β=0.07 [95%-CI, 0.05 to 0.09]), slightly smaller at the 3-year follow-up (β=0.06 [95%-CI, 0.04 to 0.08]), and attenuated at the 4-year follow-up (β=0.02 [95%-CI,-0.02 to 0.05]).

**Figure 1.**
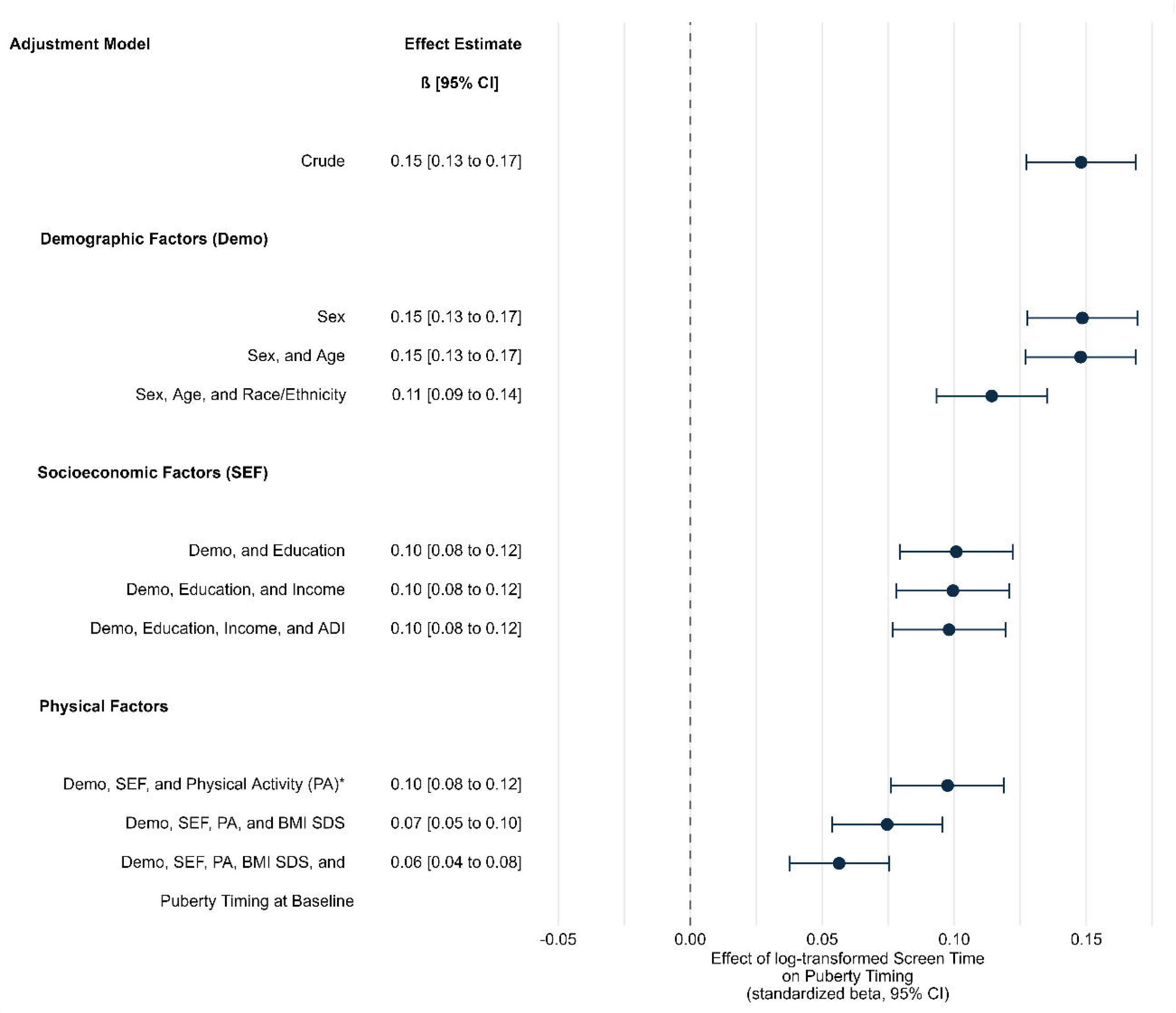
Covariate influence on the effect estimate at 2-year follow-up. Standardized betas from linear mixed models with stepwise adjustment indicate the change in pubertal timing (in SD units) per one SD increase in log-transformed screen time. Effect sizes decreased with progressive adjustment, with race/ethnicity, BMI SDS and puberty timing at baseline producing the largest changes. * The minimal sufficient adjustment set was identified using a directed acyclic graph (DAG) and included variables from the listed demographic and socioeconomic factors, as well as physical activity. It was expanded to include the BMI standard deviation score (BMI SDS) due to the association between BMI and puberty-timing [45]. For additional details, including the DAG and covariate selection procedure, see Supplementary Materials.

**Figure 2.**
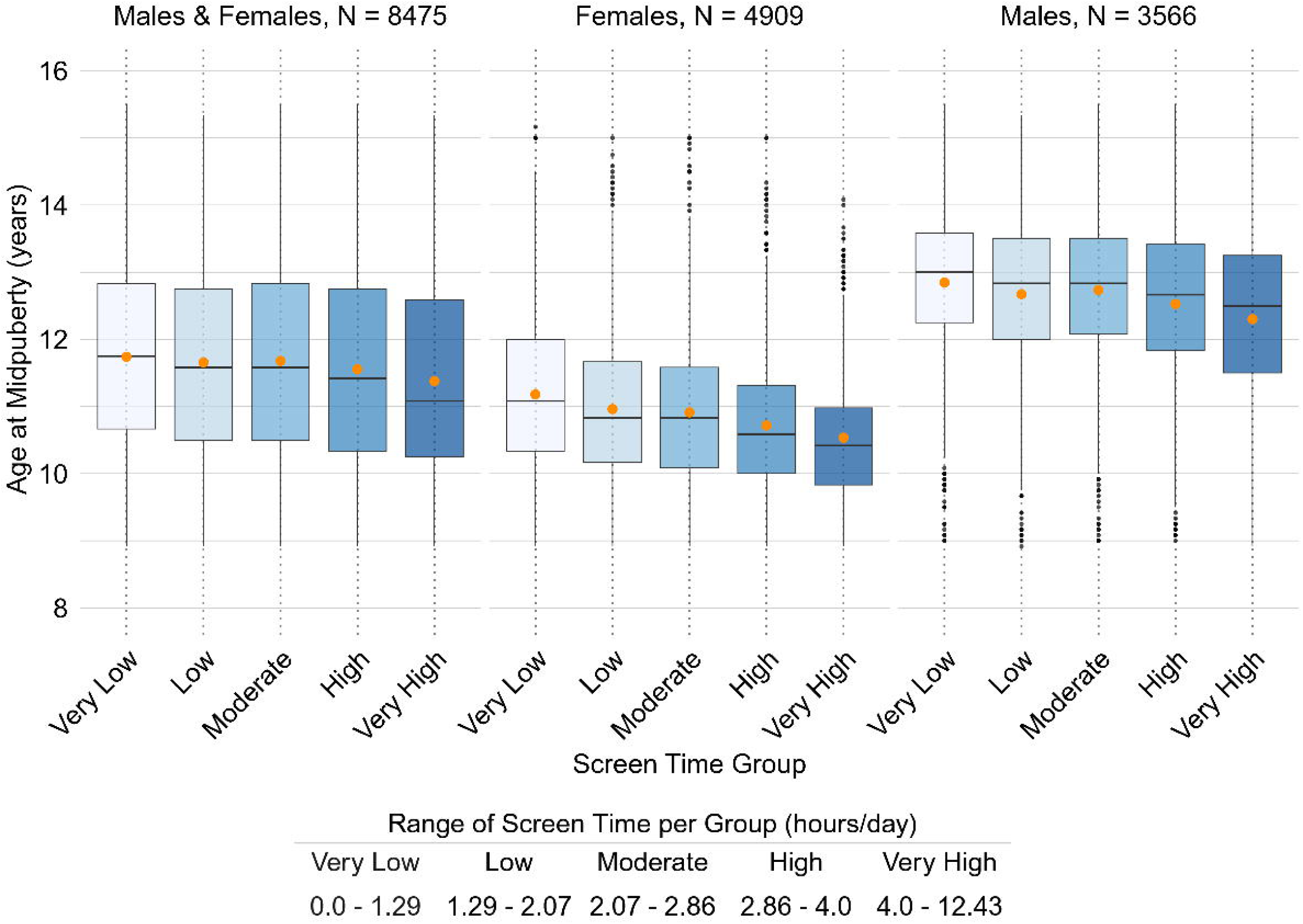
Association between screen time groups and age at mid-puberty. The boxplots show age at mid-puberty by screen time group (very low to very high), stratified by sex. Orange dots indicate means, black lines medians, boxes the IQR, and whiskers 1.5×IQR. Outliers are plotted individually. The screen time range panel was created and the final composite figure assembled using BioRender.

### Age at Mid-Puberty

Mean age at reaching mid-puberty decreased progressively across screen-time groups, ranging from 0.99 months younger in the low group to 4.35 months younger in the very high group compared with the very low group (crude differences, Figure 3, Supplementary Table S12). Adjusted linear mixed models confirmed this association, showing that higher screen time was linked to younger age at mid-puberty: B=-1.22 months [95%-CI, -2.07 to -0.38) for the low group, B=-0.84 months [95%-CI, -1.70 to 0.02] for the moderate group, B=-1.76 months [95%-CI, -2.64 to -0.89] for the high group, and B=-2.47 months [95%-CI, -3.38 to -1.56] for the very high group (Supplementary Table S12).

**Figure 3.**
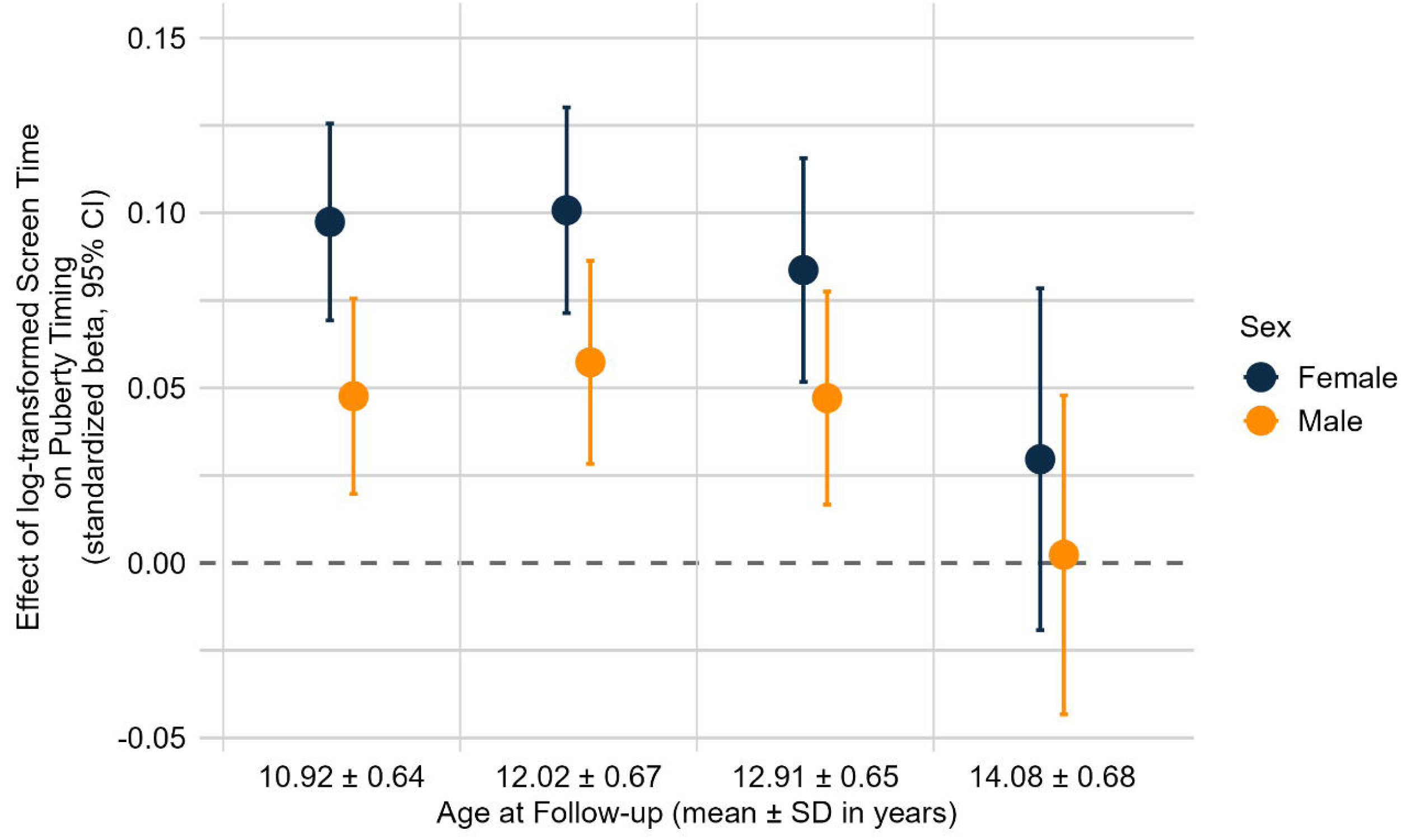
Sex-stratified effects of screen time on puberty timing. This figure presents standardized beta coefficients (95% CI) for the association between log-transformed baseline screen time and pubertal timing across follow-up years, stratified by sex. Linear mixed models adjusted for age, race/ethnicity, socioeconomic factors, physical activity, and BMI SDS. Effects were consistently larger in girls, largest at year 2.

### Results of Sensitivity Analysis

Results were consistent across all sensitivity analyses, including models using youth-reported screen time, untransformed screen time, and complete case analyses, confirming the robustness of the primary finding of an association between screen time and pubertal timing (Supplementary Table S11, Supplementary Figure S12).

### Results of Additional Analyses

We further examined the specificity of the association between baseline screen time and pubertal development. First, sequential adjustment of individual covariates at the 2-year follow-up showed that race/ethnicity, BMI SDS, and baseline pubertal timing attenuated the effect most strongly, whereas adjustments for age, parental education, income, Area Deprivation Index, and physical activity had only a minor influence (Figure 2, Supplementary Table S13).

Second, sex-stratified adjusted linear mixed models indicated that baseline screen time was associated with earlier pubertal timing in both sexes, with stronger associations observed in females than males (Figure 3, Supplementary Table S14 and S15). The strongest effects were observed in females at both the 1- and 2-year follow-ups (ß=0.10 [95%-CI, 0.07 to 0.13]) and in males at the 2-year follow-up (ß=0.06 [95%-CI, 0.03 to 0.09]).

Third, associations between baseline log-transformed screen time and puberty tempo from baseline to 2-year follow-up were small (β=0.04 [95%-CI, 0.02 to 0.06], Supplementary Table[S16).

Fourth, a small positive association between baseline youth-reported social media use and pubertal timing was observed at year 1 (β=0.02 [95%-CI, 0.01 to 0.04]). For years 2–4, effect estimates were smaller with their 95%-confidence intervals including zero (Supplementary Table S17, Supplementary Figure S13).

## Discussion

This study of the ABCD cohort (N=10786) investigated the relationship between parent-reported baseline screen time (mean age 9.91±0.63 years) and subsequent pubertal timing up to a follow-up age of 14.08±0.68 years. Screen time at baseline was consistently associated with accelerated pubertal development at follow-up. After adjusting for relevant confounders, children in the very high screen time group reached mid-puberty 2.47 months earlier compared with peers in the very low screen time group. The associations remained consistent across multiple models and covariate adjustments, and is comparable in order of magnitude to approximately a decade of secular change in the timing of thelarche reported in epidemiologic studies [6], which may be meaningful at the population level given the high prevalence of screen exposure.

Our findings align with a recent meta-analysis reporting an association between screen exposure and early puberty [18], although most included studies were cross-sectional and therefore unable to address directionality. Among the longitudinal studies in the meta-analysis, one had a small sample size and was conducted retrospectively [24], whereas the other assessed screen exposure only in terms of internet and computer use, assumed the reverse temporal direction of the association, and adjusted for only a limited number of covariates [25]. Using a longitudinal design, we found that higher screen time in late childhood was associated with more advanced pubertal maturation at subsequent follow-ups. The persistence of this association after adjustment for baseline pubertal status suggests that screen exposure in late childhood is linked to subsequent pubertal trajectories across adolescence, consistent with the temporal ordering of exposure and outcome in our study. The meta-analysis also suggested possible sex specific patterns but did not detect significant associations in sex stratified analyses [18]. In our study, effects were evident in both sexes and were nearly twice as large in girls as in boys (Figure 1, Supplementary Table S14).

Several pathomechanisms have been proposed to explain how screen exposure might act as an endocrine disruptor influencing pubertal timing. One hypothesis, supported by experimental studies in rats, suggests that exposure to blue light (short wavelength 450 to 470 nm) emitted by screen devices suppresses melatonin, which normally inhibits kisspeptin release, thereby facilitating pubertal progression [15-17, 26-28]. Other proposed mechanisms involve sedentary behavior and obesity, both well-documented consequences of excessive screen use [2, 29]. Higher body weight is a well-established risk factor for earlier pubertal maturation, especially in girls, potentially because leptin produced by adipose tissue stimulates kisspeptin release and thereby advances pubertal onset [30]. In our study, adjustment for physical activity and BMI attenuated but did not eliminate the association between screen time and pubertal timing, suggesting the involvement of additional mechanisms (Figure 1). Furthermore, psychosocial factors such as stress or anxiety related to social media have been discussed as potential contributors to accelerated pubertal timing [18]. In our analyses, self-reported social media use showed effects in the same direction but of smaller magnitude than overall screen time (Supplementary Figure S13), indicating that social media use alone is unlikely to be the primary driver of the observed association. However, the exact mechanism through which screen time may influence pubertal development remains unclear and warrants further investigation.

### Public Health Implications

The association here described between higher screen use and earlier pubertal onset adds to the expanding evidence on the health risks associated with screen exposure in childhood. Several studies indicate that early puberty has adverse consequences for later health trajectories, including a higher risk of psychopathology and risky behaviors [31-34], reduced cognitive functions [35], cardiovascular disease [36], and increased breast cancer risk [37], making early puberty a negative determinant of later health [38]. Thus, our study emphasizes the necessity of limiting screen time in childhood and underscores the need for continued public health efforts to reduce screen exposure. Behavioral interventions have shown promise for reducing screen time, with short-term interventions in early childhood being most effective [39, 40].

Several proposals have been made for upper limits on children’s screen time. Based on the 2001 AAP recommendation limiting television viewing to two hours per day, multiple studies adopted this cutoff when examining associations between screen time and pubertal timing [41]. A recent meta-analysis found no differences in pubertal timing between children above versus below this threshold (≤2 h/day versus ≥2 h/day) [18]. In 2016, the AAP revised its guidance, advising that children aged 2 to 5 years limit use to one hour per day, and that school-aged children follow a balanced approach without a fixed upper limit [42]. Our findings suggest a logarithmic association between screen exposure and pubertal timing (Supplementary Figures S5 and S6), with effects detectable even at lower exposure levels. This pattern suggests that the previously proposed threshold of 2 hours per day may not adequately capture the dose-response relationship, though the clinical significance of small effects at modest exposure levels remains uncertain.

### Limitations

Our study has several limitations. First, screen time was assessed retrospectively through parent reports rather than objective measures such as app-based screen tracking. The lack of objective screen time measurement raises the possibility of exposure misclassification, and only moderate agreement was observed between parent- and self-reported screen time (Supplementary Results). However, it should be noted that our main findings did not differ when analyses were conducted using self-reported instead of parent-reported screen time (Supplementary Figure S12). Second, factors that influence light exposure, such as viewing distance and device type, were not obtained, which limits our ability to evaluate specific mechanistic pathways, including the hypothesis that blue light exposure contributes to earlier puberty timing. Third, pubertal status was based on parent report rather than clinical Tanner staging by trained medical personnel. Although previous studies indicate moderate agreement between these methods [20], outcome misclassification cannot be excluded. Fourth, given the prolonged follow-up period, adjustment at baseline alone cannot account for time-varying confounding, so residual confounding remains possible. At the same time, adjusting for time-updated covariates could introduce overadjustment if these variables are influenced by prior screen exposure.

## Conclusion

In this longitudinal study of 10786 participants, higher screen time in late childhood was associated with earlier pubertal maturation relative to same-age, same-sex peers. After adjusting for relevant covariates, children with very high screen time reached mid-puberty approximately 2.47 months earlier than those with minimal screen time. The associations were present in both sexes but more pronounced in girls. Given the potential negative consequences of early puberty for mental and physical health, our findings highlight an additional facet of the risks associated with high screen use and underscore the importance of keeping screen time as low as possible.

## Abbreviations

95%-CI: 95% confidence interval
ABCD Study: Adolescent Brain and Cognitive Development Study
BMI: Body Mass Index
N: Number of Participants
PDS: Pubertal Timing Development Scale
SD: Standard Deviation
SDS: Standard Deviation Score

## Authors Contributions Statement

*Luise Bläschke* conceptualized and designed the study, interpreted the data, drafted the initial manuscript, carried out the statistical analysis, and critically reviewed and revised the manuscript for important intellectual content.

*Franka Edith Weisner* interpreted the data and critically reviewed and revised the manuscript for important intellectual content.

*Anke Hinney* interpreted the data, supervised the study, and critically reviewed and revised the manuscript for important intellectual content.

*Triinu Peters* interpreted the data and critically reviewed and revised the manuscript for important intellectual content.

*Börge Schmidt* interpreted the data and critically reviewed and revised the manuscript for important intellectual content.

*Raphael Hirtz* conceptualized and designed the study, interpreted the data, supervised the study, and critically reviewed and revised the manuscript for important intellectual content.

*Lars Dinkelbach* conceptualized and designed the study, interpreted the data, supervised the study, and critically reviewed and revised the manuscript for important intellectual content.

All authors approved the final manuscript as submitted and agree to be accountable for all aspects of the work.

## Data sharing

This study used data from the Adolescent Brain Cognitive Development (ABCD) Study, a National Institutes of Health (NIH) research project. ABCD data are available via the National Institute of Mental Health Data Archive (NDA) under controlled access. Researchers can request data access via the NDA data application process (https://nda.nih.gov/abcd). Data access is granted after approval to ensure compliance with ethical guidelines and participant confidentiality. For further information, please visit the official ABCD study websites [43].

## Additional Information

Lars Dinkelbach was supported by a fellowship from the University Medicine Essen Clinician Scientist Academy (UMEA) of the Medical Faculty of the University Duisburg-Essen (supported by the German Research Foundation (DFG)) to LD (FU 356/12-2). Artificial intelligence (ChatGPT version 5.1, Open AI) was used between October 1 and December 6, 2025 to improve the manuscript’s language style (spelling, grammar, phrasing, and overall writing quality) of the Introduction, Methods, Results, and Discussion. Luise Bläschke is responsible for the accuracy and integrity of the final content.

## Ethics

The ABCD study protocol received approval from the central Institutional Review Board at the University of California, San Diego, as well as from local Institutional Review Boards at selected study sites (Clark et al., 2018). Approval for the use of the data in the present study was granted by the Ethics Committee of the University of Duisburg Essen (No. 24-12040-BO).

