## Supplementary Material for "Screen Time and Puberty Timing: Findings from the Adolescent Brain Cognitive Development Study"

Supplementary Methods 1 – ABCD sample analyses

Study Sample

Supplementary Figure S1. Exclusion Criteria and Final Sample Composition

Supplementary Table S1. Cohort Characteristics of the Study Sample, Male and Female Participants at Baseline

Supplementary Table S2. Characteristics of Included and Excluded Participants

Exposure: Screentime

Supplementary Figure S2. Average Daily Screen Time at Baseline

Outcome: Puberty Timing

Supplementary Figure S3. Relative Puberty Timing, PDS Sum Score (Parent Reported) and Age of Female Participants by Event

Supplementary Figure S4- Puberty Timing, PDS Sum Score (Parent Reported) and Age of Male Participants by Event

Linear Mixed Models – Assumptions

Supplementary Figure S5. Effect of Untransformed Screen Time on Puberty Timing.

Supplementary Figure S6. Effect of Log-Transformed Screen Time on Puberty Timing

Supplementary Table S3. Akaike Information Criterion Comparison of Linear Mixed Models for Different Screen Time Transformations

Supplementary Figure S7. Quantile-Quantile (Q–Q)

Supplementary Table S4. Variance Inflation Factors Across Follow-Up Years (Males and Females Combined)

Covariates and Missingness

Supplementary Figure S8. DAG to Identify a Minimal Sufficient Adjustment Set.

Supplementary Table S5. Comparison of Subjects With and Without Available Family Income Information

Supplementary Table S6. Comparison of Participants With and Without Available Parental Education Information

Supplementary Table S7. Comparison of Subjects With and Without Available Area Deprivation Index Information

Supplementary Table S8. Comparison of Participants With and Without Available Physical Activity Information

Supplementary Table S9. Observed and Imputed Total Family Income Distributions

Supplementary Table S10. Observed and Imputed Highest Parental Education Distributions

Supplementary Figure S9. Observed and Imputed Physical Activity Distributions

Supplementary Figure S10. Observed and Imputed Area Deprivation Index (ADI; Percentile) Distributions

Sensitivity Analysis

Supplementary Figure S11. Comparison of Screen Time by Reporter

Supplementary Results

Supplementary Table S11. Sensitivity Analyses: Robustness of Screen Time Associations with Puberty Timing

Supplementary Table S12. Association Between Screen Time Group and Age at Mid-Puberty

Supplementary Figure S12. Sensitivity Analysis: Comparison of Estimated Effects of Parent- vs. Youth-Reported Screen Time

Supplementary Table S13. Further Analysis: Influence of Covariates on the Effect Estimate of Baseline Screen Time on Puberty Timing

Supplementary Table S14. Further Analyses: Sex-Specific Associations between Screen Time and Puberty Timing

Supplementary Table S15. Further Analyses: Sex-Specific Associations Between Screen Time Group and Age at Midpuberty

Supplementary Table S16. Further Analyses: Screen Time and Puberty Tempo

Supplementary Table S17. Further Analyses: Social Media Screen Time and Puberty Timing

Supplementary Figure S13. Additional Analysis: Comparison of Estimated Effects of Parent-Reported Screentime and Youth-Reported Social Media Use.

Supplementary References

**Supplementary Methods 1 - ABCD sample analyses**

Participant numbers with N<10 are not reported in accordance with the ABCD Study data reporting guidelines.

**Study Sample**

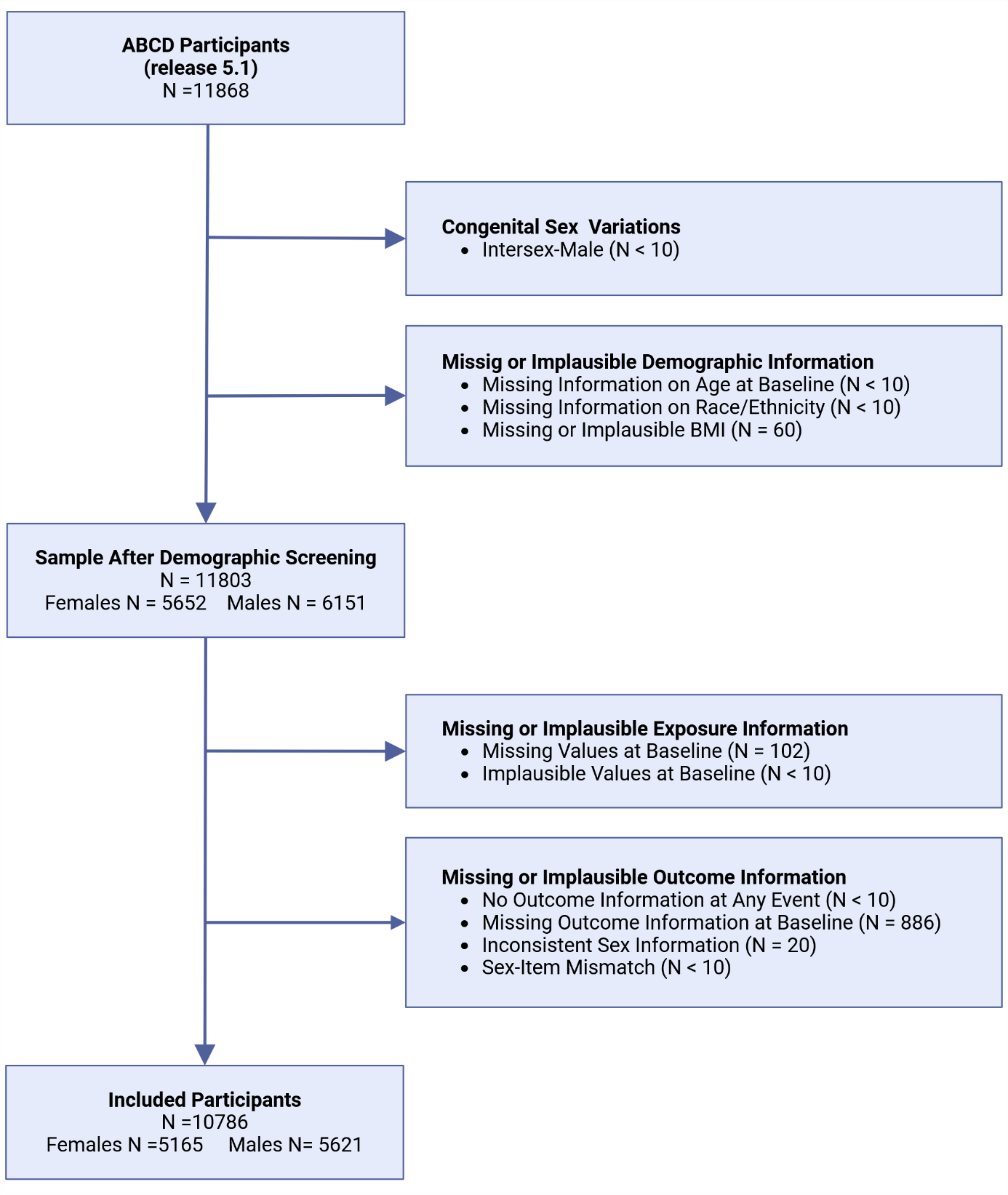

**Supplementary Figure S1. Exclusion Criteria and Final Sample Composition.**
The figure illustrates the exclusion process used to form the final study sample and describes its composition. Figure created with BioRender.com.

Abbreviations: N = number of participants; BMI = body mass index.

| **Supplementary Table S1. Cohort Characteristics of the Study Sample, Male and Female Participants at Baseline** | | | | |
| --- | --- | --- | --- | --- |
|  | **Total Sample** | **Male Participants** | | **Female Participants** |
| **Characteristics** | **(N=10786)** | **(N=5621)** | | **(N=5165)** |
| **Sex, N (% of Study Sample)** | | | | |
|  | | | 5621 (52.11) | 5165 (47.89) |
| **Age in Years, mean (SD)** | | | | |
|  | 9.91 (0.63) | 9.93 (0.63) | | 9.90 (0.62) |
| **Race/Ethnicity, N (%)** | | | | |
| White | 5839 (54.13) | 3095 (55.06) | | 2744 (53.13) |
| Black | 1446 (13.41) | 709 (12.61) | | 737 (14.27) |
| Hispanic | 2135 (19.79) | 1116 (19.85) | | 1019 (19.73) |
| Asian | 229 (2.12) | 110 (1.96) | | 119 (2.30) |
| Other | 1137 (10.54) | 591 (10.51) | | 546 (10.57) |
| **Body Mass Index** | | | | |
| kg/m², mean (SD) | 18.77 (4.16) | 18.68 (4.04) | | 18.87 (4.28) |
| SDS^a^, mean (SD) | 0.41 (1.15) | 0.44 (1.15) | | 0.38 (1.15) |
| **Highest Parental Education, N^a^ (%)** | | | | |
| Less than high school | 451 (4.18) | 223 (3.97) | | 228 (4.41) |
| High school diploma or GED | 923 (8.56) | 469 (8.34) | | 454 (8.79) |
| Some college education | 1347 (12.49) | 721 (12.83) | | 626 (12.12) |
| Associate degree | 1407 (13.04) | 733 (13.04) | | 674 (13.05) |
| Bachelor's degree | 2824 (26.18) | 1491 (26.53) | | 1333 (25.81) |
| Postgraduate education | 3834 (35.55) | 223 (3.970) | | 228 (4.41) |
| **Total Family Income, N^b^ (%)** | | | | |
| < $5000 | 404 (3.75) | 199 (3.54) | | 205 (3.97) |
| $5,000 - $11,999 | 409 (3.79) | 209 (3.72) | | 200 (3.87) |
| $12,000 - $15,999 | 256 (2.37) | 129 (2.29) | | 127 (2.46) |
| $16,000 - $24,999 | 541 (5.02) | 294 (5.23) | | 247 (4.78) |
| $25,000 - $34,999 | 629 (5.83) | 316 (5.62) | | 313 (6.06) |
| $35,000 - $49,999 | 935 (8.67) | 483 (8.59) | | 452 (8.75) |
| $50,000 - $74,999 | 1452 (13.46) | 771 (13.72) | | 681 (13.18) |
| $75,000 - $99,999 | 1569 (14.55) | 812 (14.45) | | 757 (14.66) |
| $100,000 - $199,999 | 3311 (30.70) | 1742 (30.99) | | 1569 (30.38) |
| > $200,000 | 1280 (11.87) | 666 (11.85) | | 614 (11.89) |

Abbreviations:

N	Number of Participants

SD	Standard Deviation

SDS	Standard deviation score

GED	General Educational Development

^a^	BMI SDS, adjusted for age and sex using the 2000 CDC growth reference data for U.S. children and adolescents [1], represents the deviation of an individual’s BMI from the mean BMI of the reference population.

^b^	The reported values include imputed data; details on the information used, including selection criteria, conditions, and the imputation procedure, are provided in the Supplementary Methods Covariates and Missingness*.*

| **Supplementary Table S2. Characteristics of Included and Excluded Participants** | | | |
| --- | --- | --- | --- |
|  | **Included Participants** | **Excluded Participants** | **p values ^a^**  **t-test/Χ²-test** |
|  | (N=10786) | (N=1082) |  |
| **Sex, N (%)** | | | |
| Male | 5621 (52.11) | 567 (52.40) | < 0.001 |
| Female | 5165 (47.89) | 512 (47.32) |  |
| **Age in Years, mean (SD)** | | | |
|  | 9.9 (0.6) | 9.9 (0.6) | 0.209 |
| **Race/Ethnicity, N (%)** | | | |
| White | 5839 (54.13) | 334 (30.87) | < 0.001 |
| Black | 1446 (13.41) | 338 (31.24) |  |
| Hispanic | 2135 (19.79) | 275 (25.42) |  |
| Asian | 229 (2.12) | 23 (2.13) |  |
| Other | 1137 (10.54) | 111 (10.26) |  |
| NA | 0 (0) | <10 (<0.92) |  |
| **Highest Parental Education, N^b^ (%)** | | | |
| Less than high school | 449 (4.16) | 144 (13.31) | < 0.001 |
| High school diploma or GED | 920 (8.53) | 212 (19.59) |  |
| Some college education | 1346 (12.48) | 159 (14.70) |  |
| Associate degree | 1406 (13.04) | 163 (15.06) |  |
| Bachelor's degree | 2823 (26.17) | 190 (17.56) |  |
| Postgraduate education | 3832 (35.53) | 210 (19.41) |  |
| NA | 10 (0.09) | <10 (<0.92) |  |
| **Family Income, N (%)** | | | |
| < $5,000 | 322 (2.99) | 95 (8.78) | < 0.001 |
| $5,000 - $11,999 | 346 (3.21) | 75 (6.93) |  |
| $12,000 - $15,999 | 225 (2.09) | 48 (4.44) |  |
| $16,000 - $24,999 | 461 (4.27) | 62 (5.73) |  |
| $25,000 - $34,999 | 567 (5.26) | 87 (8.04) |  |
| $35,000 - $49,999 | 841 (7.80) | 93 (8.60) |  |
| $50,000 - $74,999 | 1365 (12.66) | 133 (12.29) |  |
| $75,000 - $99,999 | 1477 (13.69) | 93 (8.60) |  |
| $100,000 - $199,999 | 3155 (29.25) | 156 (14.42) |  |
| > $200,000 | 1207 (11.19) | 43 (3.97) |  |
| “Refuse to answer” | 417 (3.87) | 94 (8.69) |  |
| “Don't know” | 403 (3.74) | 101 (9.33) |  |
| NA | 0 (0) | <10 (<0.92) |  |

Abbreviations:

N	Number of Participants

SD	Standard deviation

NA	Not Available

^a^	Categorical variables, including sex, race/ethnicity, and family income class, were analyzed using Chi-squared tests. Age and Area Deprivation Index, as continuous variables, were assessed with a t-test.

^b^	Highest Parental Education was aggregated from the original categories without any imputation. See Supplementary Methods Covariates and Missingness for a detailed description of the aggregation procedure.

*Exposure : Screen Time at Baseline*

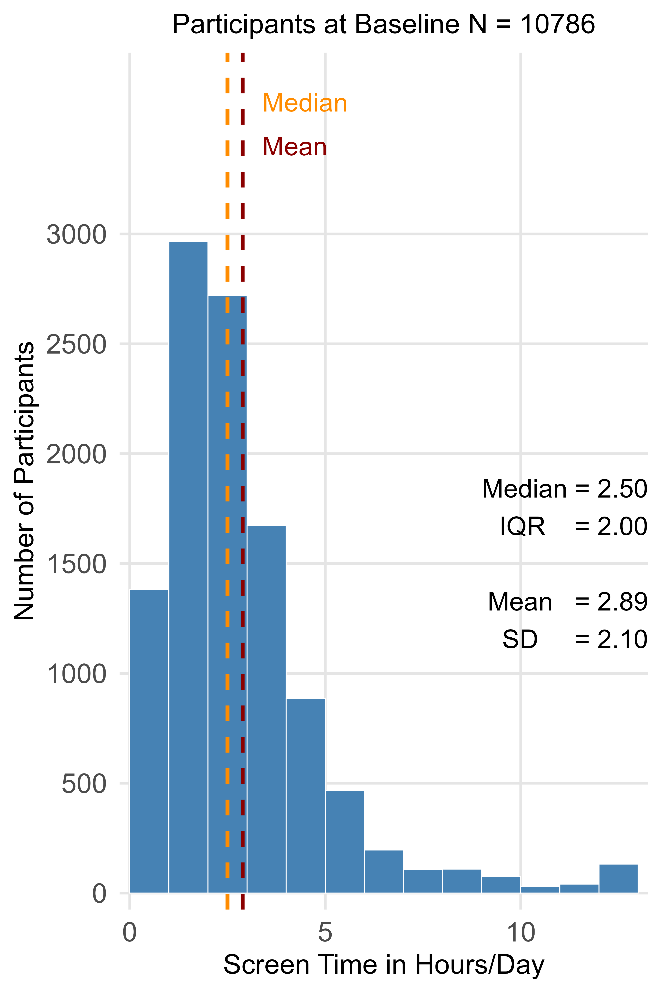

**Supplementary Figure S2**. **Average Daily Screen Time at Baseline.** Distribution of the reported daily screen time (see Methods).

*Outcome: Puberty Timing*

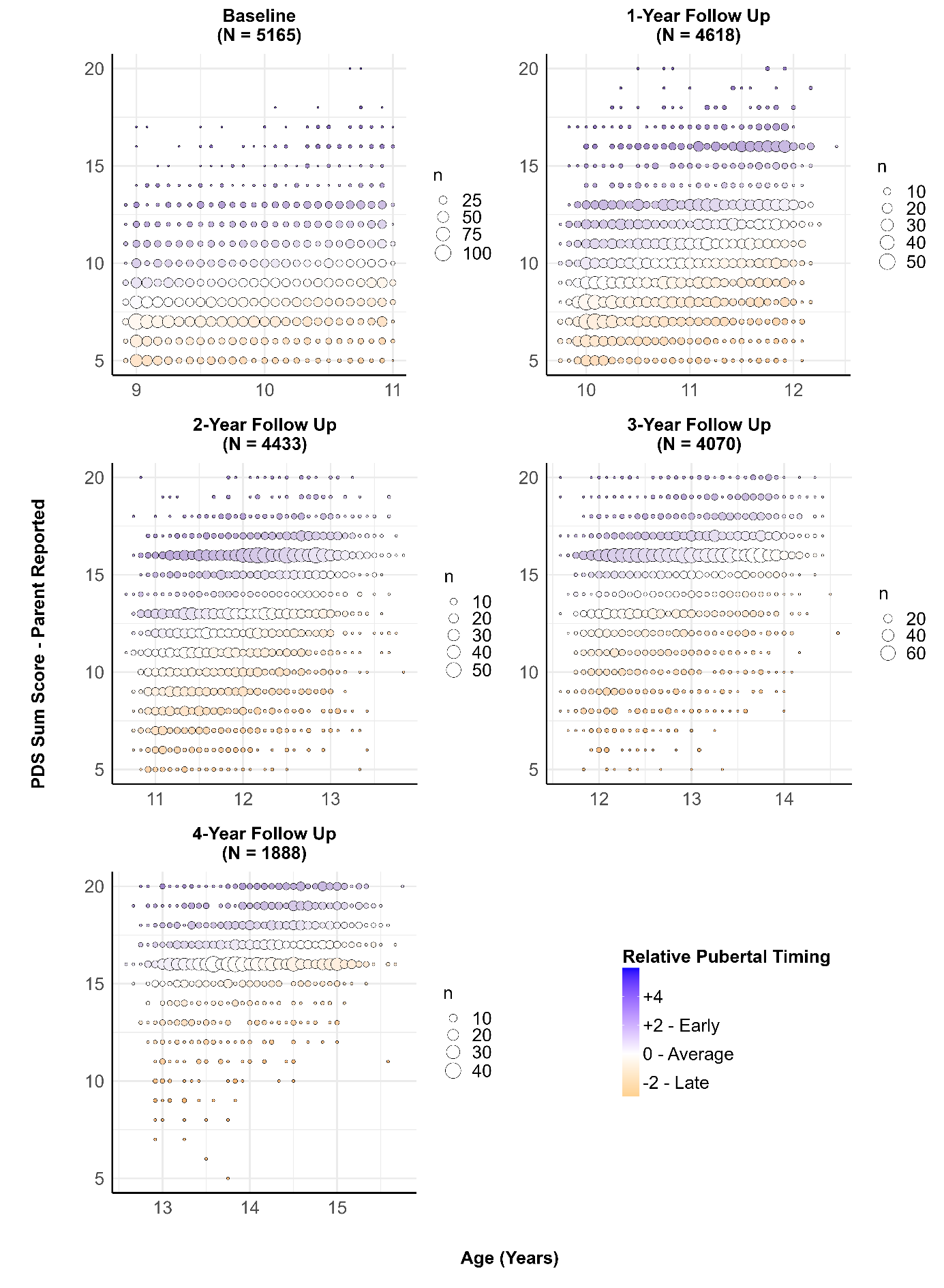
**Supplementary Figure S3. Relative Puberty Timing, PDS Sum Score (Parent Reported), and Age of Female Participants by Event.** Pubertal timing was operationalized by performing a regression analysis of the Pubertal Development Scale (PDS) sum scores on age within each sex group. The standardized residuals obtained from these regressions were subsequently used as indicators of individual pubertal timing. The figure illustrates the relationship between puberty timing, age, and PDS total score for each assessment time point in female participants. Blue indicates positive standardized residuals, representing earlier-than-average puberty timing. Age (in years) is plotted on the x-axis, and the PDS total score, ranging from 5 to 20, is shown on the y-axis. The size of each circle is proportional to the number of participants with the same combination of PDS score and age.

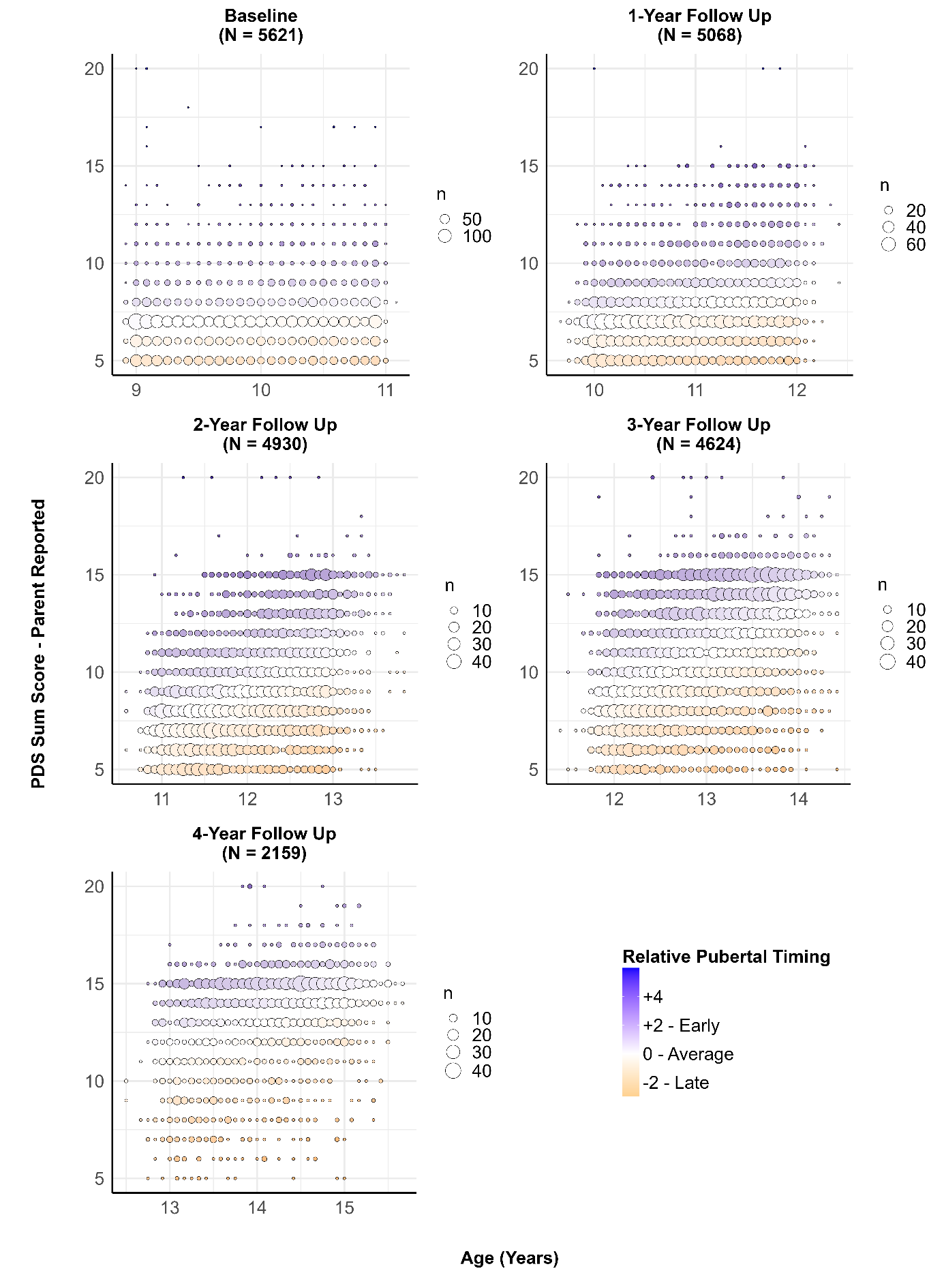

**Supplementary Figure S4. Puberty Timing, PDS Sum Score (Parent Reported), and Age of Male Participants by Event.** Pubertal timing was operationalized by performing a regression analysis of the Pubertal Development Scale (PDS) sum scores on age within each sex group. The standardized residuals obtained from these regressions were subsequently used as indicators of individual pubertal timing. The figure illustrates the relationship between puberty timing, age, and PDS total score for each assessment time point in male participants. Blue indicates positive standardized residuals, representing earlier-than-average puberty timing. Age (in years) is plotted on the x-axis, and the PDS total score, ranging from 5 to 20, is shown on the y-axis. The size of each circle is proportional to the number of participants with the same combination of PDS score and age.

*Linear Mixed Models – Assumptions*

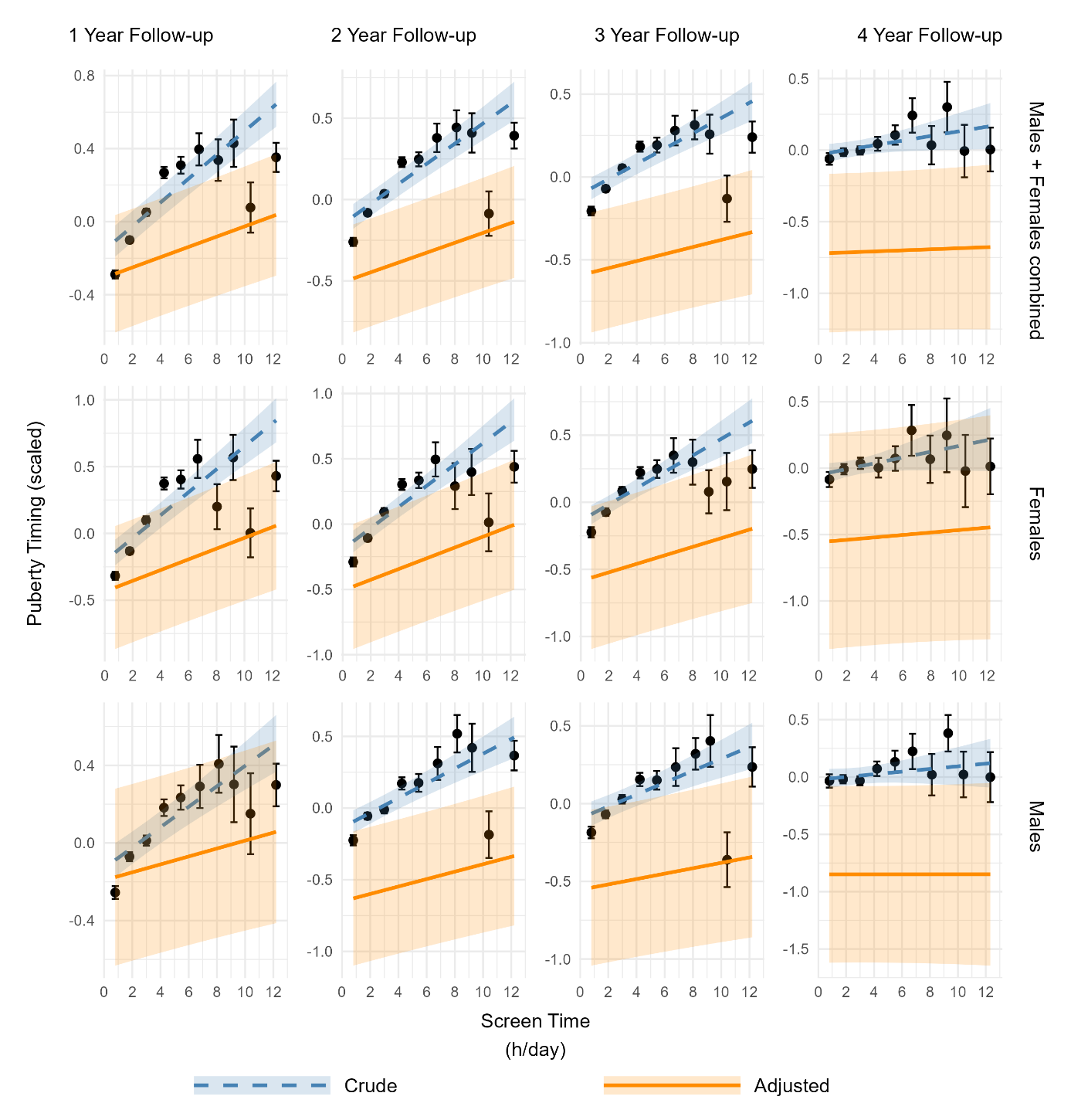

**Supplementary Figure S5. Effect of Untransformed Screen Time on Puberty Timing.** To visualize the linear effects of screen time on puberty timing, the screen time was divided into 10 equally spaced intervals. For each interval, the black dot represents the mean of the standardized outcome, the standard error of the mean (SEM) is shown as vertical error bars, and the midpoint of the interval is used as the x-value for the dots. The ‘Crude Model’ (blue dashed line with 95% confidence band) shows the linear effect of standardized screen time on standardized puberty timing, including random intercepts for site and family nested within site. The ‘Adjusted Model’ (orange solid line with 95% confidence band) additionally incorporates baseline covariates: age, race/ethnicity, highest parental education, total family income, Area Deprivation Index, weekly physical activity, and BMI-SDS, while keeping the same random effects structure. For models stratified by sex (males or females), sex was not included. Without transformation, the association between screen time and pubertal timing deviated from linearity, showing larger increases in pubertal timing at lower screen time levels and smaller, attenuated increases at higher levels, consistent with a logarithmic pattern.

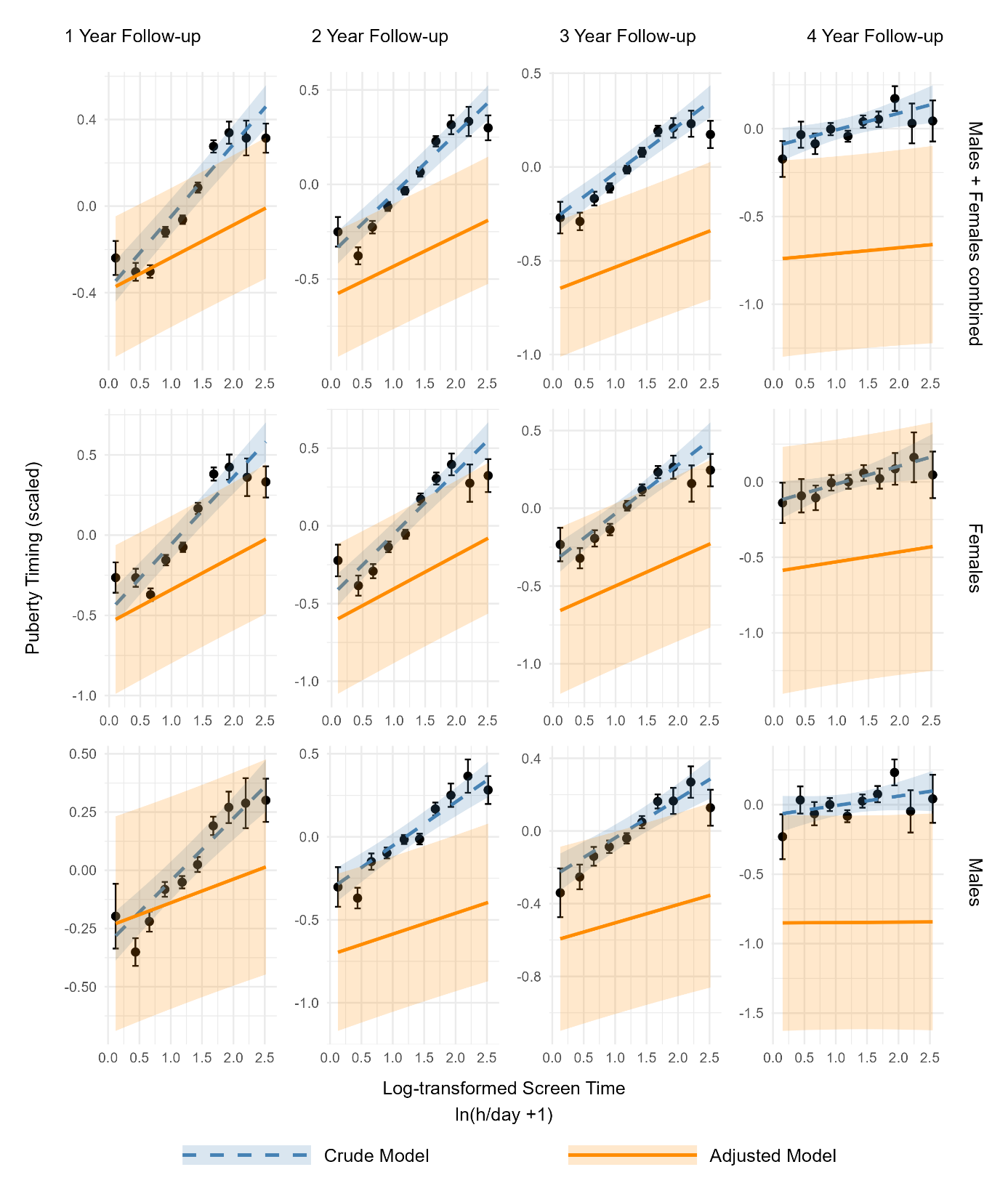

**Supplementary Figure S6. Effect of Log-Transformed Screen Time on Puberty Timing.** To visualize the linear effects of log-transformed screen time on puberty timing, the log-transformed screen time was divided into 10 equally spaced intervals. For each interval, the black dot represents the mean of the standardized outcome, the standard error of the mean (SEM) is shown as vertical error bars, and the midpoint of the interval is used as the x-value for the dots. The ‘Crude Model’ (blue dashed line with 95% confidence band) shows the linear effect of standardized log-transformed screen time on standardized puberty timing, including random intercepts for site and family nested within site. The ‘Adjusted Model’ (orange solid line with 95% confidence band) additionally incorporates baseline covariates: age, race/ethnicity, highest parental education, total family income, Area Deprivation Index, weekly physical activity, and BMI-SDS, while keeping the same random effects structure. For models stratified by sex (males or females), sex was not included. After log-transformation, the effect of screen time on puberty timing roughly followed a linear pattern.

*Model fit*: To compare model fit across different screen time transformations, the Akaike Information Criterion (AIC) was computed for the adjusted linear mixed models, separately for each follow-up year. For easier comparison, the difference in AIC (ΔAIC) was derived relative to the AIC of the model with untransformed screen time. Assuming that a ΔAIC ≥2 indicates a meaningful difference, the models using log-transformed screen time show meaningful differences compared with untransformed screen time for follow-up years 1 to 3 (Supplementary Table S3) [16]. Among all transformations tested, the log transformation consistently yielded the best model fit; accordingly, log-transformed screen time was used in the main analyses.

| **Supplementary Table S3.** **Akaike Information Criterion Comparison of Linear Mixed Models for Different Screen Time Transformations** | | | | | |
| --- | --- | --- | --- | --- | --- |
| **Type of Screen Time Transformation** | | **Untransformed** | **Square Root** | **Exponential** | **Logarithmic** |
| **1 Year Follow-up** | | | | | |
|  | AIC | 25372.31 | 25361.50 | 25405.95 | 25358.74 |
|  | Δ AIC | Ref | 10.81 | - 32.64 | 13.57 |
| **2 Year Follow-up** | | | | | |
|  | AIC | 24952.92 | 24942.63 | 24985.81 | 24939.68 |
|  | Δ AIC | Ref | 10.29 | - 32.89 | 13.25 |
| **3 Year Follow-up** | | | | | |
|  | AIC | 23649.57 | 23640.74 | 23665.12 | 23637.13 |
|  | Δ AIC | Ref | 8.83 | - 15.55 | 12.44 |
| **4 Year Follow-up** | | | | | |
|  | AIC | 11290.34 | 11289.84 | 11289.85 | 11289.69 |
|  | Δ AIC | Ref | 0.50 | -0.49 | 0.65 |

Abbreviations:

AIC	Akaike Information Criterion

Δ AIC	Difference in Akaike Information Criterion (AIC), calculated by subtracting the AIC of the model with untransformed screen time from the AIC of the respective model.

*Normality of residuals:* To assess the normality of residuals in the linear mixed-effects models for males and females combined, QQ plots were generated for log-transformed screen time across all follow-up years to compare the standardized residuals from the sample with the theoretical quantiles. The visually observed deviations from normality were considered negligible given the large sample size of the study.

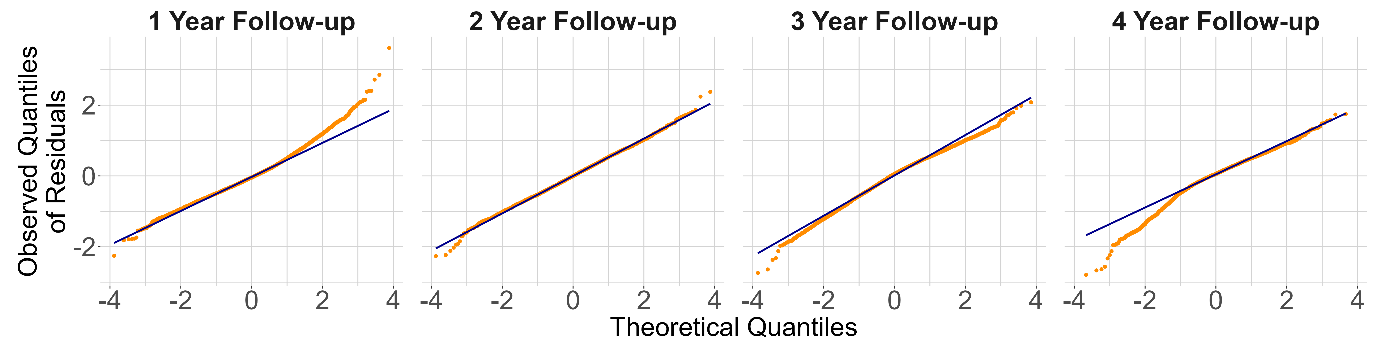
**Supplementary Figure S7. Quantile-Quantile (Q–Q)** plots were used to assess the normality of residuals from the adjusted models analyzing log-transformed screen time across the follow-up periods, with males and females analyzed together.

*Multicollinearity*: To assess multicollinearity among predictors, generalized variance inflation factors (GVIFs) were calculated. The calculations were based on linear models for males and females combined, included only the fixed effects, and used log-transformed screen time as a predictor, while random effects were not considered. To allow comparison across categorical variables with differing numbers of levels, the GVIFs were scaled using the transformation GVIF^(1/(2*df)), and the squared values of these adjusted GVIFs (i.e., GVIF^(1/df)) were interpreted. Assuming that a squared adjusted GVIF exceeding 4–5 indicated potentially problematic multicollinearity [15], none of the covariates in the present models reached this threshold, indicating that problematic multicollinearity was unlikely (Supplementary Table S4).

| **Supplementary Table S4. Variance Inflation Factors Across Follow-Up Years (Males and Females Combined)** | | | | | | | | | |
| --- | --- | --- | --- | --- | --- | --- | --- | --- | --- |
|  | | **1 Year Follow-Up** | | **2 Year Follow-Up** | | **3 Year Follow-Up** | | **4 Year Follow-Up** | |
| **Variable** | | GVIF | GVIF^1/df^ | GVIF | GVIF^1/df^ | GVIF | GVIF^1/df^ | GVIF | GVIF^1/df^ |
|  | Screen Time^a^ | 1.15 | 1.15 | 1.15 | 1.15 | 1.16 | 1.16 | 1.16 | 1.16 |
|  | Age | 1.02 | 1.02 | 1.02 | 1.02 | 1.02 | 1.02 | 1.02 | 1.02 |
|  | Sex | 1.01 | 1.01 | 1.01 | 1.01 | 1.02 | 1.02 | 1.02 | 1.02 |
|  | Race/Ethnicity | 1.64 | 1.13 | 1.65 | 1.13 | 1.64 | 1.13 | 1.57 | 1.12 |
|  | Highest Parental Education | 2.32 | 1.18 | 2.36 | 1.19 | 2.30 | 1.18 | 2.21 | 1.17 |
|  | Family Income | 2.64 | 1.11 | 2.69 | 1.12 | 2.59 | 1.11 | 2.51 | 1.11 |
|  | Area Deprivation Index | 1.55 | 1.55 | 1.55 | 1.55 | 1.52 | 1.52 | 1.47 | 1.47 |
|  | Physical Activity | 1.04 | 1.04 | 1.04 | 1.04 | 1.04 | 1.04 | 1.05 | 1.05 |
|  | BMI - SDS | 1.11 | 1.11 | 1.10 | 1.10 | 1.10 | 1.10 | 1.10 | 1.10 |

Abbreviation:

BMI-SDS	Body Mass Index – Standard Deviation Scores

GVIF	Generalized variance inflation factors

GVIF^1/df^	squared adjusted GVIF

*Covariates and Missingness*

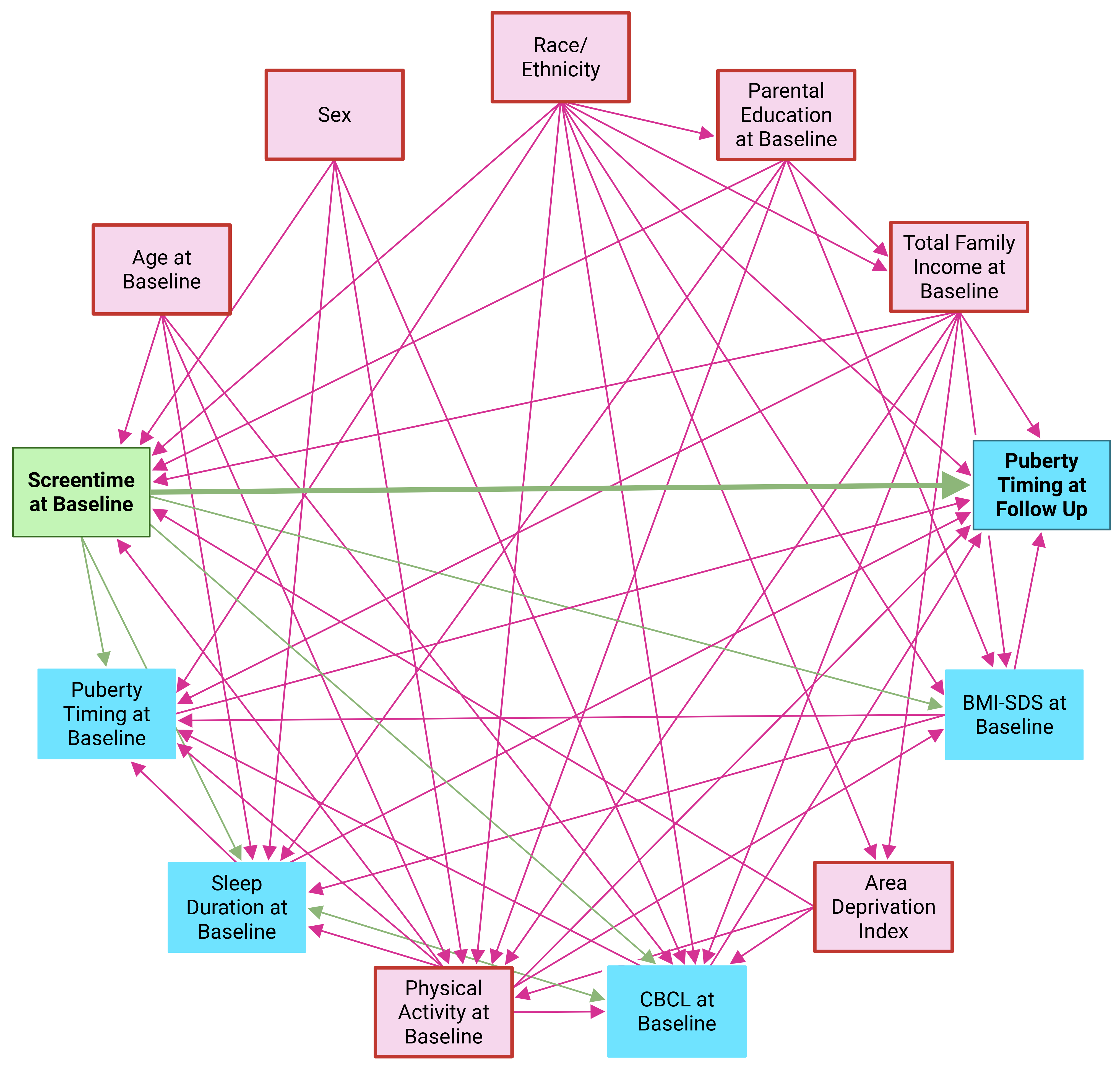

**Supplementary Figure S8. DAG to Identify a Minimal Sufficient Adjustment Set.** The online tool DAGitty was used to create a directed acyclic graph (DAG) to examine the relationships between potential confounders, the exposure (screen time at baseline), and the outcome (puberty timing at follow-up), and to identify the minimal set of variables for adjustment to estimate the causal effect of the exposure on the outcome [2]. The exposure variable (in bold) is displayed with a green background, while the outcome variable (in bold) and its exclusive ancestors appear with a blue background. Shared ancestors of the exposure and outcome variables are shown with a red background. Biasing paths are represented by red arrows, and causal effects by green arrows. To identify a minimal sufficient adjustment set, we specified a causal pathway in which baseline screen exposure precedes pubertal development during follow-up, while acknowledging that earlier unmeasured pubertal trajectories cannot be fully excluded. Baseline pubertal timing was therefore treated as a potential mediator or proxy for prior maturation rather than a confounder and was not included in the primary adjustment set, which aimed to estimate the total association of baseline screen time with subsequent pubertal timing. A literature review by Eirich et al. found a correlation between screen time and externalizing/internalizing problems in children but did not clarify the direction of this relationship [5]. We consider the influence of screen time on the Child Behavior Checklist (CBCL) as an indicator of overall psychopathology relevant, and since a bidirectional relationship would not allow for adjustment as it would result in a cyclic graph, we modeled a unidirectional influence of screen time on the CBCL at baseline [6, 7]. Based on studies reporting a bidirectional association between sleep disturbances and externalizing symptoms, and a unidirectional effect of sleep on internalizing symptoms, we modeled a bidirectional relationship between sleep and CBCL in the DAG for completeness [8]. A comparison in DAGitty showed that assuming a unidirectional path from sleep to CBCL would not have altered the minimal sufficient adjustment set. Given these assumptions, the minimal sufficient adjustment set for estimating the total effect of screen time on puberty timing at follow-up includes the following variables: Age at Baseline, Family Income at Baseline, Parental Education at Baseline, Physical Activity at Baseline, Race/Ethnicity, Residential Environment, and Sex. In addition to the variables of the minimal sufficient adjustment set, we decided to correct for BMI SDS at baseline, as literature has shown that BMI is associated with pubertal development [9, 10].

*BMI*

To account for body weight, baseline BMI Standard Deviation Scores (BMI-SDS) were calculated. Weight and height measurements were converted to kilograms and centimeters, respectively, using the measurements package (v1.5.1). BMI-SDS values were derived from the CDC growth reference charts [11] with the childsds package (v0.9.11), taking each participant’s age and sex into account. BMI-SDS values could not be calculated for 15 participants due to missing or implausible information on weight or height. Assuming that BMI-SDS values below -4 or above +8 reflect implausible extreme values in the underlying height or weight data [8], the height or weight measurements of 45 additional participants were deemed implausible. Based on these criteria, a total of N (%) = 60 (0.51%) participants were excluded from the analysis.

*Missingness*

Participants with missing or implausible baseline demographic values (age, race/ethnicity, or BMI), missing baseline exposure data (“Screen Time at Baseline”), or missing outcome data (“Puberty Timing at Follow-up”) were excluded. Outcome exclusions included cases where the parent-reported PDS never contained evaluable information, baseline data were missing, or there was a mismatch between sex reported in the PDS, the demographic questionnaire, or the sex-specific items. Applying these criteria resulted in a final sample of 10786 participants (90.88 % of the ABCD study sample). Excluded participants were more frequently from ethnic minority groups and had parents with lower levels of highest education and lower total family income, compared with included participants (Supplementary Table S2). To prevent a substantial reduction of the sample due to missing covariate data, missing values were imputed using multiple imputations for Total Family Income (N= 820, 7.60 %), Highest Parental Education (N= 10, <1 %), Area Deprivation Index (N= 789, 7.32 %), and Physical Activity (N= 1653, 15.33 %).

*Total Family Income*

In the Parent Demographic Survey, total combined family income during the past 12 months was assessed using a 10-point ordinal scale (see Supplementary Table S1 for income categories). For this covariate, only baseline values were included. Income information was unavailable for N=1017 participants.

| **Supplementary Table S5. Comparison of Subjects With and Without Available Family Income Information** | | | |
| --- | --- | --- | --- |
|  | **Available** | **Missing** | **p values ^a^**  **t-test/Χ²-test** |
|  | (N=10851) | (N=1017) |  |
| **Age in Years, mean (SD)** | | | |
|  | 9.91 (0.62) | 9.93 (0.62) | 0.549 |
| **Sex, N (%)** | | | |
| Male | 5642 (52.0) | 546 (53.7) | 0.176 |
| Female | 5207 (47.99) | 470 (46.2) |  |
| **Race/Ethnicity, N (%)** | | | |
| White | 5878 (54.17) | 295 (29.01) | <0.001 |
| Black | 1511 (13.92) | 273 (26.84) |  |
| Hispanic | 2103 (19.38) | 307 (30.19) |  |
| Asian | 217 (2.00) | 35 (3.44) |  |
| Other | 1141 (10.52) | 107 (10.52) |  |
| NA | <10 (< 0.09) | 0 (0.00) |  |
| **Area Deprivation Index, Percentile , mean (SD)** | | | |
|  | 39.44 (26.60) | 46.69 (30.06) | <0.001 |
| **Highest Parental Education, N (%)** | | | |
| Less than high school | 448 (4.13) | 145 (14.26) | <0.001 |
| High school diploma or GED | 930 (8.57) | 202 (19.86) |  |
| Some college education | 1355 (12.49) | 150 (14.75) |  |
| Associate degree | 1421 (13.10) | 148 (14.55) |  |
| Bachelor's degree | 2842 (26.19) | 171 (16.81) |  |
| Postgraduate education | 3850 (35.48) | 192 (18.88) |  |
| NA | <10 (<0.09) | <10 (<0.98) |  |

Abbreviations

N	Number

SD	Standard Deviation

^a^	Categorical variables, including sex, race/ethnicity and Highest Parental Education were analyzed using Chi-squared tests. Age and Area Deprivation Index, as continuous variables, were assessed with a t-test.

*Highest Parental Education*

The variable Highest Parental Education was derived from responses in the Parent Demographic Survey. Parents were asked to report (a) their own highest level of education completed (“What is the highest grade or level of school you have completed or the highest degree you have received?”) and (b) their partner’s highest level of education (“What is the highest grade or level of school your partner completed or highest degree they received?”). Responses were recorded on an ordinal scale ranging from 0 = Never attended/Kindergarten to 21 = Doctoral degree. Valid information was available for N=11851 (99.86%) respondents and for N=9404 (79.24%) partners. For N=9401 (79.21%), both values were available, and for N=11854 (99.88%), at least one valid response was present. Missing information was defined as responses of Refused to answer (777), Don’t know (999), or no response at all, resulting in N=14 (0.12%) cases with two invalid entries. To construct the Highest Parental Education variable, the higher of the two reported values was used if both were available; if only one was available, that value was taken. For the N=14 cases with two invalid responses, the value was treated as missing.

For the analysis, the detailed 22-level parental education variable (including imputed values for missing or invalid responses) was recoded into a 6-level categorical variable according to the following scheme:

- 0 = Less than high school (original categories 0 [Never attended / Kindergarten] to 12 [12th grade])
- 1 = High school diploma or GED (original categories 13 [High school graduate] to 14 [GED or equivalent])
- 2 = Some college, no degree (original category 15 [Some college])
- 3 = Associate degree (original categories 16 [Associate degree: Occupational] to 17 [Associate degree: Academic program])
- 4 = Bachelor’s degree (original category 18 [Bachelor’s degree, e.g., BA])
- 5 = Postgraduate education (original categories 19 [Master’s degree, e.g., MA] to 21 [Doctoral degree, e.g., PhD])

| **Supplementary Table S6. Comparison of Participants With and Without Available Parental Education Information** | | | |
| --- | --- | --- | --- |
|  | **Available** | **Missing** | **p values ^a^**  **t-test/Χ²-test** |
|  | (N=11854) | (N=14) |  |
| **Age in Years, mean (SD)** | | | |
|  | 9.91 (0.62) | 9.98 (0.68) | 0.687 |
| **Sex, N (%)** | | | |
| Male | 6178 (52.12) | NA^b^ | 0.352 |
| Female | 5673 (47.86) | NA^b^ |  |
| **Race/Ethnicity, N (%)** | | | |
| White | 6172 (52.07) | <10 (<71.43) | 0.024 |
| Black | 1779 (15.01) | <10 (<71.43) |  |
| Hispanic | 2405 (20.29) | <10 (<71.43) |  |
| Asian | 252 (2.13) | 0 (0.0) |  |
| Other | 1245 (10.50) | <10 (<71.43) |  |
| NA | <10 (<0.08) | 0 (0.00) |  |
| **Area Deprivation Index, Percentile , mean (SD)** | | | |
|  | 40.04 (26.97) | 46.55 (34.02) | 0.424 |
| **Total Family Income, N (%)** | | | |
| < $5,000 | 416(3.51) | <10 (<71.43) | < 0.001 |
| $5,000 - $11,999 | 421 (3.55) | 0 (0.00) |  |
| $12,000 - $15,999 | 272 (2.29) | <10 (<71.43) |  |
| $16,000 - $24,999 | 523 (4.41) | 0 (0.00) |  |
| $25,000 - $34,999 | 653 (5.51) | <10 (<71.43) |  |
| $35,000 - $49,999 | 932 (7.86) | <10 (<71.43) |  |
| $50,000 - $74,999 | 1498 (12.64) | 0 (0.00) |  |
| $75,000 - $99,999 | 1570 (13.24) | 0 (0.00) |  |
| $100,000 - $199,999 | 3311 (27.93) | 0 (0.00) |  |
| > $200,000 | 1250 (10.54) | 0 (0.00) |  |
| “Refuse to answer” | 503 (4.24) | <10 (<71.43) |  |
| “Don't know” | 503 (4.24) | <10 (<71.43) |  |
| NA | <10 (0.08) | 0 (0.00) |  |

Abbreviations

N	Number of participants

SD	Standard Deviation

^a^	Categorical variables, including sex, race/ethnicity and Family Income Education were analyzed using Chi-squared tests. Age and Area Deprivation Index, as a continuous variables, were assessed with a t-test.

^b^	Distribution not shown due to small sample size; see note on the ABCD Study data reporting guidelines above.

*Area Deprivation Index*

The Area Deprivation Index (ADI), expressed as a national percentile score, was used to assess neighborhood socioeconomic disadvantage [12] and was calculated based on Kind et al. and provided in the ABCD Study dataset [13]. For N=879 participants (7.41%), ADI values were missing and were imputed.

| **Supplementary Table S7. Comparison of Subjects With and Without Available Area Deprivation Index Information** | | | |
| --- | --- | --- | --- |
|  | **Available** | **Missing** | **p values ^a^**  **t-test/Χ²-test** |
|  | (N=10989) | (N=879) |  |
| **Age in Years, mean (SD)** | | | |
|  | 9.92 (0.62) | 9.88 (0.62) | 0.117 |
| **Sex, N (%)** | | | |
| Male | 5753 (52.35) | 435 (49.49) | 0.230 |
| Female | 5233 (47.62) | 444 (50.51) |  |
| **Race/Ethnicity, N (%)** | | | |
| White | 5770 (52.51) | 403 (45.85) | <0.001 |
| Black | 1579 (14.37) | 205 (23.32) |  |
| Hispanic | 2238 (20.37) | 172 (19.57) |  |
| Asian | 239 (2.17) | 13 (1.48) |  |
| Other | 1162 (10.57) | 86 (9.78) |  |
| NA | <10 (<0.09) | 0 (0.00) |  |
| **Highest Parental Education, N (%)** | | | |
| Less than high school | 530(4.82) | 63 (7.17) | 0.004 |
| High school diploma or GED | 1036 (9.43) | 96 (10.92) |  |
| Some college education | 1384 (12.59) | 121 (13.77) |  |
| Associate degree | 1463 (13.31) | 106 (12.06) |  |
| Bachelor's degree | 2806 (25.53) | 207 (23.55) |  |
| Postgraduate education | 3759 (34.21) | 283 (32.20) |  |
| NA | 11 (0.10) | <10 (<1.14) |  |
| **Total Family Income, N (%)** | | | |
| < $5,000 | 373 (3.39) | 44 (5.01) | <0.001 |
| $5,000 - $11,999 | 391 (3.56) | 30 (3.41) |  |
| $12,000 - $15,999 | 250 (2.28) | 23 (2.62) |  |
| $16,000 - $24,999 | 475 (4.32) | 48 (5.46) |  |
| $25,000 - $34,999 | 603 (5.49) | 51 (5.80) |  |
| $35,000 - $49,999 | 877 (7.98) | 57 (6.48) |  |
| $50,000 - $74,999 | 1407 (12.80) | 91 (10.35) |  |
| $75,000 - $99,999 | 1484 (13.50) | 86 (9.78) |  |
| $100,000 - $199,999 | 3077 (28.00) | 234 (26.62) |  |
| > $200,000 | 1137 (10.35) | 113 (12.86) |  |
| “Refuse to answer” | 462 (4.20) | 49 (5.57) |  |
| “Don't know” | 451 (4.10) | 53 (6.03) |  |
| NA | <10 (<0.09) | 0 (0.00) |  |

Abbreviations

N	Number of Participants

^a^	Categorical variables, including sex, race/ethnicity, highest parental education and total family income were analyzed using Chi-squared tests. Age, as a continuous variable, was assessed with a t-test.

*Physical Activity*

Baseline physical activity of participants was assessed using the Parent Sports and Activities Involvement Questionnaire (SAIQ), which included questions on 23 different sports. Parents reported, for each sport, the number of months per year their child participated during the most active period (1 = 1 month to 12 = 12 months), the number of days per week (1–7 = 1–7 days per week, 8 = once every 2 weeks, 9 = one day per month, 10 = less than one day per month), and the duration of each session in minutes (1 = less than 30 minutes, 2 = 30 minutes, 3 = 45 minutes, 4 = 60 minutes, 5 = 90 minutes, 6 = 120 minutes, 7 = 150 minutes, 8 = 180 minutes, 9 = greater than 3 hours). Responses coded as 999 (“Don’t know”) were treated as missing. Days per week and minutes per session were recoded to approximate numerical values (days per week: 1 = 1, 2 = 2, …, 7 = 7, 8 = 0.5, 9 = 0.25, 10 = 0.1, 999 = NA; minutes per session: 1 = 15, 2 = 30, 3 = 45, 4 = 60, 5 = 90, 6 = 120, 7 = 150, 8 = 180, 9 = 210, 999 = NA), and months per year were capped at 12. For each sport, weekly minutes of activity were calculated as the product of months, days per week, and minutes per session divided by 52, provided that none of the three values were missing. These weekly minutes were then summed across all sports to derive a total weekly physical activity score per participant. For participants with missing information on all sports, physical activity was imputed (N=1995 [16.81 %]).

| **Supplementary Table S8. Comparison of Participants With and Without Available Physical Activity Information** | | | |
| --- | --- | --- | --- |
|  | **Available** | **Missing** | **p values ^a^**  **t-test/Χ²-test** |
|  | (N=9873) | (N=1995) |  |
| **Age in Years, mean (SD)** | | | |
|  | 9.93 (0.63) | 9.85 (0.62) | <0.001 |
| **Sex, N (%)** | | | |
| Male | 5215 (52.82) | 973 (48.77) | 0.003 |
| Female | 4655 (47.15) | 1022 (51.23) |  |
| **Race/Ethnicity, N (%)** | | | |
| White | 5580 (56.52) | 593 (29.72) | <0.001 |
| Black | 1186 (12.01) | 598 (29.97) |  |
| Hispanic | 1872 (18.96) | 538 (26.97) |  |
| Asian | 222 (2.25) | 30 (1.50) |  |
| Other | 1012 (10.25) | 236 (11.83) |  |
| NA | <10 (<0.10) | 0 (0.00) |  |
| **Area Deprivation Index, Percentile , mean (SD)** | | | |
|  | 36.84 (25.56) | 56.14 (28.16) | <0.001 |
| **Highest Parental Education, N^b^ (%)** | | | |
| Less than high school | 324 (3.28) | 269 (13.48) | <0.001 |
| High school diploma or GED | 718 (7.27) | 414 (20.75) |  |
| Some college education | 1105 (11.19) | 400 (20.05) |  |
| Associate degree | 1190 (12.05) | 379 (19.00) |  |
| Bachelor's degree | 2701 (27.36) | 312 (15.64) |  |
| Postgraduate education | 3825 (38.74) | 217 (10.88) |  |
| NA | 10 (0.10) | <10 (<0.50) |  |
| **Family Income, N (%)** | | | |
| < $5,000 | 235 (2.38) | 182 (9.12) | <0.001 |
| $5,000 - $11,999 | 272 (2.75) | 149 (7.47) |  |
| $12,000 - $15,999 | 178 (1.80) | 95 (4.76) |  |
| $16,000 - $24,999 | 339 (3.43) | 184 (9.22) |  |
| $25,000 - $34,999 | 469 (4.75) | 185 (9.27) |  |
| $35,000 - $49,999 | 717 (7.26) | 217 (10.88) |  |
| $50,000 - $74,999 | 1230 (12.46) | 268 (13.43) |  |
| $75,000 - $99,999 | 1389 (14.07) | 181 (9.07) |  |
| $100,000 - $199,999 | 3120 (31.60) | 191 (9.57) |  |
| > $200,000 | 1222 (12.38) | 28 (1.40) |  |
| “Refuse to answer” | 383 (3.88) | 128 (6.42) |  |
| “Don't know” | 319 (3.23) | 185 (9.27) |  |
| NA | 0 (0.00) | <10 (<0.50) |  |

Abbreviations

N	Number of participants

SD	Standard deviation

NA	Not available

^a^	Categorical variables, including sex, race/ethnicity, highest parental education and total family income were analyzed using Chi-squared tests. Age, as a continuous variable, was assessed with a t-test.

*Imputation of unavailable Information*

Missing data can be categorized as MCAR (completely random, independent of observed or unobserved variables), MAR (dependent only on observed variables), and MNAR (dependent on unobserved variables). Supplementary Tables S5–S8 indicate that missing information for Total Family Income, Highest Parental Education, the Area Deprivation Index, and Physical Activity appears to be associated with demographic variables. Since distinguishing between MAR and MNAR was not possible, Multiple Imputation by Chained Equations (MICE; mice package version 3.17., [14]) was applied under the assumption of MAR. However, some degree of selection bias and residual uncertainty due to potential deviations from the MAR assumption cannot be excluded.

Factor variables, including Total Family Income and Highest Parental Education, were imputed using polytomous regression (polyreg) with appropriate predictors: Total Family Income was predicted by site-ID, Race/Ethnicity, and Highest Parental Education, while Highest Parental Education was predicted by site-ID, Race/Ethnicity, and Total Family Income. Numerical variables, including the Area Deprivation Index and Physical Activity, were imputed using predictive mean matching (pmm); the Area Deprivation Index was predicted by site-ID, Race/Ethnicity, and Total Family Income, and Physical Activity was predicted by site-ID, sex, Race/Ethnicity, BMI, BMI z-score, and Age at baseline.

| **Supplementary Table S9. Observed and Imputed Total Family Income Distributions** | | |
| --- | --- | --- |
|  | **Observed** | **Imputed^a^** |
|  | (N=10851) | (N=1017) |
| **Total Family Income, N (%)** | | |
| < $5,000 | 417 (3.84) | 104 (10.23) |
| $5,000 - $11,999 | 421 (3.88) | 86 (8.46) |
| $12,000 - $15,999 | 273 (2.52) | 51 (5.01) |
| $16,000 - $24,999 | 523 (4.82) | 94 (9.24) |
| $25,000 - $34,999 | 654 (6.03) | 92 (9.05) |
| $35,000 - $49,999 | 934 (8.61) | 109 (10.72) |
| $50,000 - $74,999 | 1498 (13.81) | 117 (11.50) |
| $75,000 - $99,999 | 1570 (14.47) | 108 (10.62) |
| $100,000 - $199,999 | 3311 (30.51) | 176 (17.31) |
| > $200,000 | 1250 (11.52) | 80 (7.87) |

Abbreviations

N	Number of participants

^a^	Values in this column represent imputed data

| **Supplementary Table S10. Observed and Imputed Highest Parental Education Distributions** | | |
| --- | --- | --- |
|  | **Observed** | **Imputed^a^** |
|  | (N=11854) | (N=14) |
| **Highest Parental Education, N (%)** | | |
| Less than high school | 593 (5.00) | 2 (14.29) |
| High school diploma or GED | 1132 (9.55) | 3 (21.43) |
| Some college education | 1505 (12.70) | 3 (21.43) |
| Associate degree | 1569 (13.24) | 3 (21.43) |
| Bachelor's degree | 3013 (25.42) | 1 (7.14) |
| Postgraduate education | 4042 (34.10) | 2 (14.29) |

Abbreviations

N	Number of participants

GED	General Educational Development certificate

^a^	Values in this column represent imputed data and do not allow any conclusions regarding the identities of the study participants; values with N<10 are therefore reported

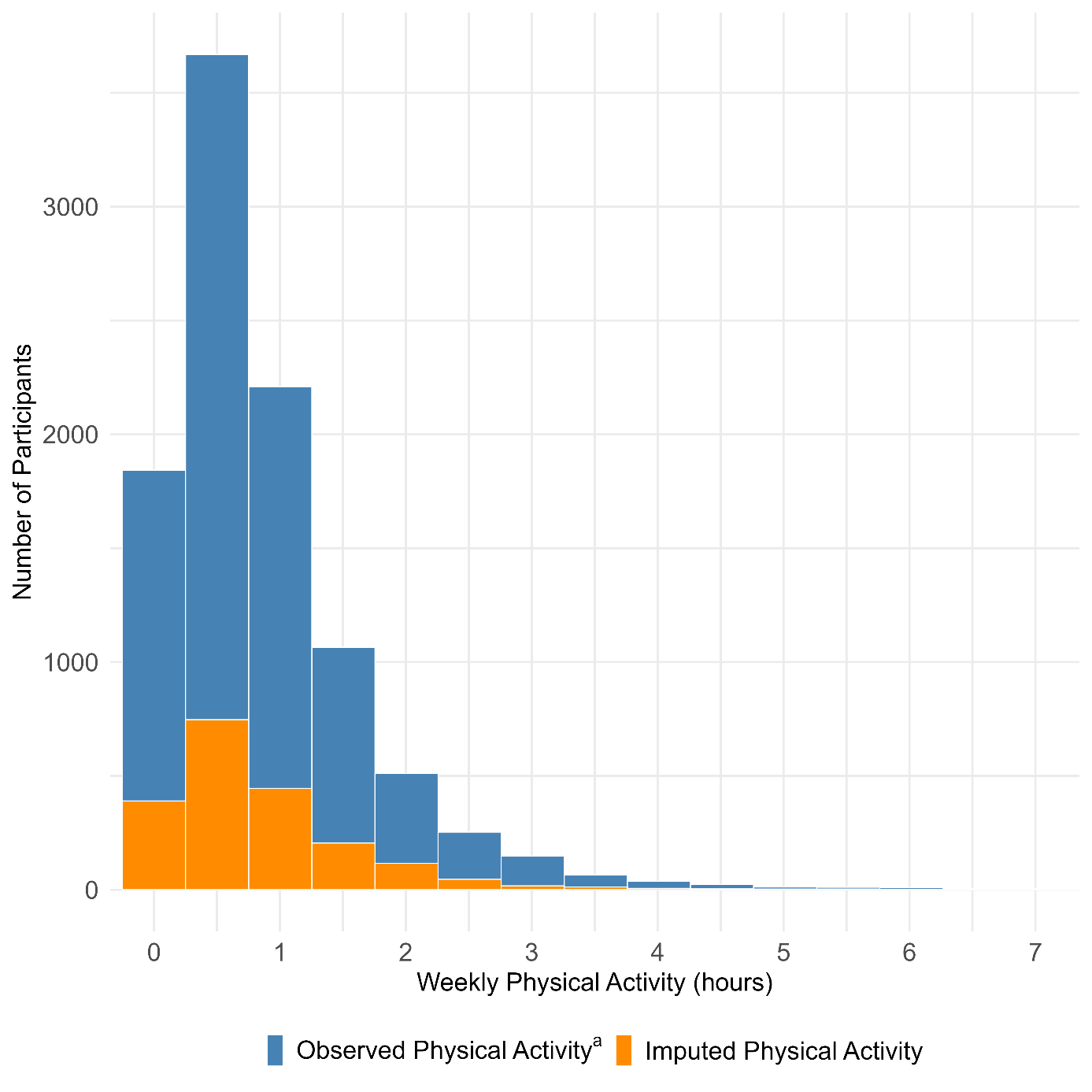

**Supplementary Figure S9. Observed and Imputed Physical Activity Distributions**

Abbreviations

^a^	Due to the small number of participants with more than 7 hours of observed weekly physical activity, which would not have been visually discernible in the histogram, the x-axis was limited to 7 hours. As a result, N = 10 participants with activity levels ranging from 7.21 to 14.5 hours per week are not displayed

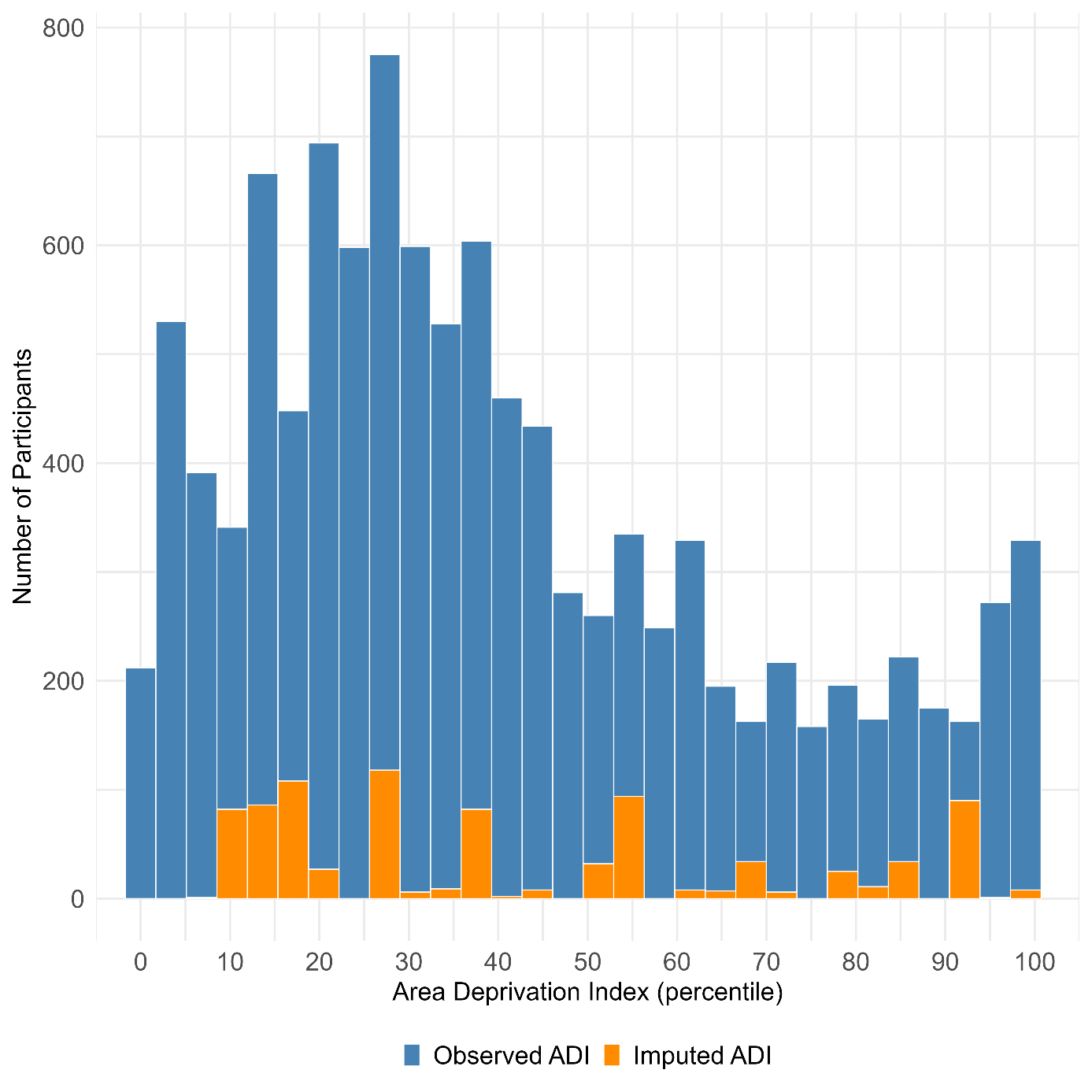

**Supplementary Figure S10. Observed and Imputed Area Deprivation Index (ADI; Percentile) Distributions**

*Sensitivity Analyses*

*Youth Reported Screentime:* Youth Reported Screen Time was calculated based on participants’ responses in the Youth Screen Time Survey. Participants were asked to indicate how many hours they typically spend on a weekday or weekend day with a) “Watch TV shows or movies?”, b) “Watch videos (such as YouTube)?”, c) “Play video games on a computer, console, phone or other device (Xbox, PlayStation, iPad)?”, d) “Text on a cell phone, tablet, or computer (e.g., Chat, WhatsApp, etc.)?”, e) “How many hours do you: Visit social networking sites like Facebook, Twitter, Instagram, etc.?”, f) “How many hours do you: Video chat (Skype, Facetime, etc.)?”. The responses were recorded on an ordinal scale: “0 = None; 0.25 = < 30 minutes; 0.5 = 30 minutes; 1 = 1 hour; 2 = 2 hours; 3 = 3 hours; 4 = 4+ hours // Example: 1½ hours would be coded as 1 hour, rather than 2 hours.” A sum score was calculated for both weekdays and weekend days, and these values are provided in the ABCD data. To allow comparison with the screen time values from the parent report, the sum scores were used as an approximation of the typical screen time in hours per day. A daily score was calculated using a weighted average under the assumption of 5 weekdays and 2 weekend days. In total, 10766 participants could be included in the sensitivity analysis based on the availability of youth reported screen time. Analogous to the parent reported screen time values, the youth reported screen time values were winsorized at the 99th percentile (>14.50 h/day, N=106).

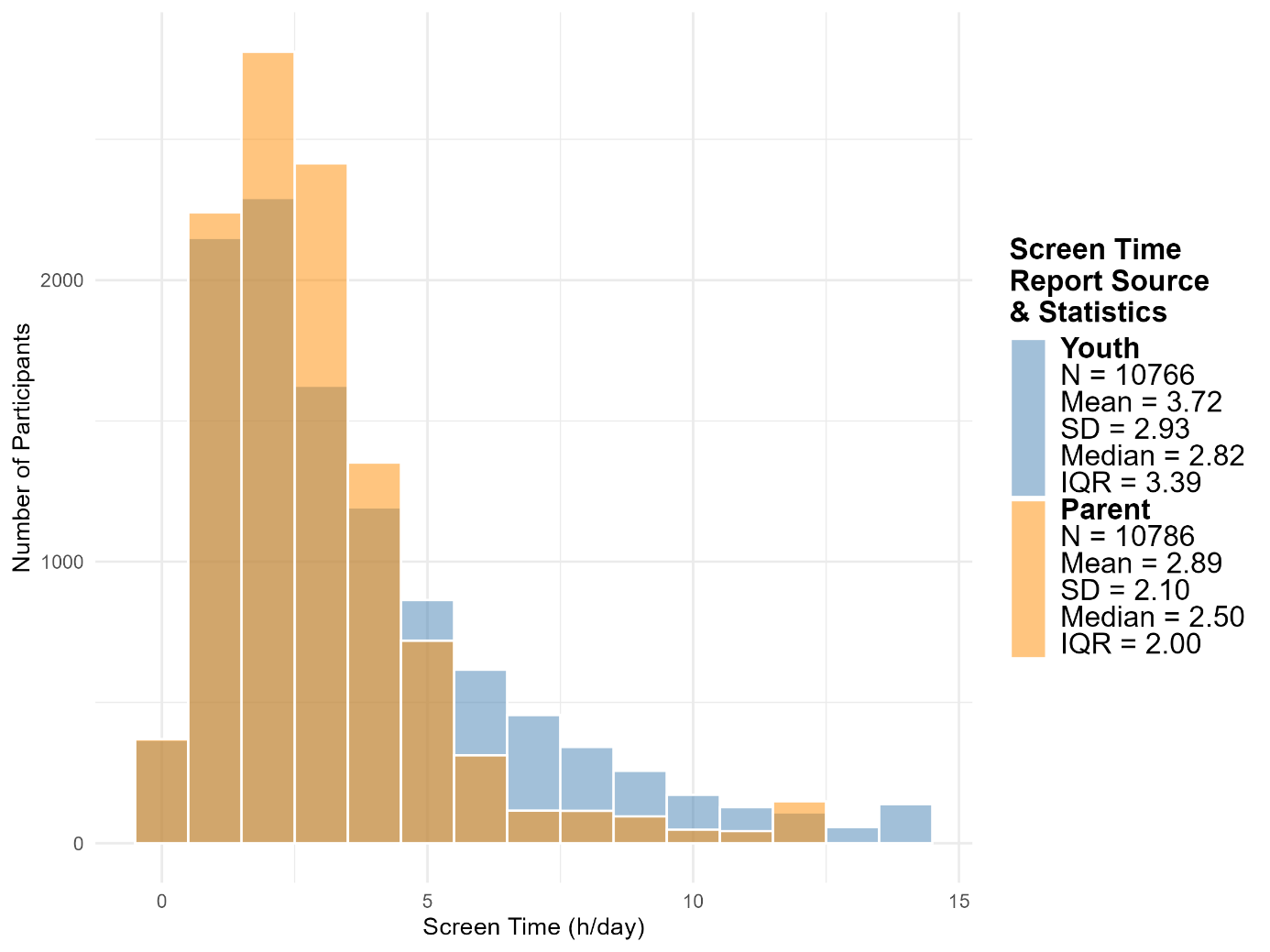

**Supplementary Figure S11. Comparison of Screen Time by Reporter**. The x-axis shows daily screen time and the y-axis represents the number of participants. Blue bars indicate youth-reported screen time, while yellow bars represent parent-reported screen time. Screen time values for both reporters were winsorized at the 99th percentile. Youth-reported screen time was higher than parent-reported screen time, with a mean (SD) of 3.72 (2.93) versus 2.89 (2.10), and a median (IQR) of 2.82 (3.39) versus 2.50 (2.0), respectively.

To investigate the association between screen time at baseline and puberty timing at follow-up, youth-reported screen time was log-transformed. Adjusted linear mixed models were run for both sexes combined and for each follow-up year 1–4 (see Supplementary Table S11, Supplementary Figure S12). The agreement between parent- and self-reported screen time was assessed using screen time groups. For N=8475 participants who reached mid-puberty during the observation period from baseline to the 4-year follow-up, youth-reported screen time was assessed as described above. Based on these values, participants were categorized into five equally sized screen time groups: very low (N=1695, range of screen time 0 -1.29 h/day), low (N=1695, 1.29 - 2.07 h/day), moderate (N=1695, 2.07 – 2.86 h/day), high (N=1695, 2.86 – 4.00 h/day), and very high (N=1695, 4.00 –12.43 h/day). Spearman’s rank correlation analysis between the parent- and youth-reported screen time group revealed a moderate positive association (ρ=0.41, p<0.001).

*Untransformed Screen Time:* In the main analysis of the effect of screen time at baseline on puberty timing at follow-up, screen time values were log-transformed to improve the normality of the residuals and the linearity of the effects in the mixed models. A sensitivity analysis using untransformed values for males and females combined was conducted to assess the robustness of the results (Supplementary Table S11).

*Exclusion of Imputed Data:* In the main analysis of the effect of screen time at baseline on puberty timing at follow-up and the effect of screen time group on age at mid-puberty, missing values for the covariates were imputed. To assess the robustness of the results, a sensitivity analysis was conducted in which all participants with imputed data were excluded, which led to 7930 participants with complete baseline data for screen time, age, sex, race/ethnicity, highest parental education, total family income, BMI-SDS, Area Deprivation Index, and physical activity. Results of adjusted linear mixed models are shown in Supplementary Table S11.

*Additional Analyses*

*Analysis by sex*: Analyses were conducted separately by sex to account for potential sex-specific associations between screen time and puberty timing (Supplementary Table S14) or between screen time and age at mid-puberty (Supplementary Table S15).

*Puberty Tempo*: Puberty tempo was defined as the annualized change in Pubertal Development Scale (PDS) sum score between baseline and the 2 year follow up, computed as the difference in PDS sum scores (2 year minus baseline) divided by the elapsed time in years. Higher values indicate faster pubertal progression.

*Social Media Screen Time*: Social media screen time at baseline was calculated based on participants’ responses to the Youth Screen Time Survey. Participants reported, separately for typical weekdays and weekend days, ‘How many hours do you: Visit social networking sites like Facebook, Twitter, Instagram, etc.?’. Responses on the ordinal scale, as described above in the Youth Reported Screen Time section, were treated as approximations of actual daily usage in hours. A weighted daily average was then calculated, assuming 5 weekdays and 2 weekend days, and the resulting values were logarithmically transformed. A total of N=10763 (99.79 % of study sample) participants were included in the analysis of the association between social media screen time and puberty timing (Supplementary Figure S13, Supplementary Table S17).

*Supplementary Results*

| **Supplementary Table S11. Sensitivity Analyses: Robustness of Screen Time Associations with Puberty Timing** | | | | | | | | | | | | | | | | | |
| --- | --- | --- | --- | --- | --- | --- | --- | --- | --- | --- | --- | --- | --- | --- | --- | --- | --- |
|  | | **Logarithmic transformed Screen Time – Parent Report**  **Primary Analysis** | | | | **Untransformed Screen Time – Parent Report** | | | | **Logarithmic transformed Screen Time – Youth Report** | | | | **Logarithmic Screen Time – Parent Report – Participants with Imputed Values excluded** | | | |
| **Follow-Up** | | N | β | 95% CI | p | N | β | 95% CI | p | N | β | 95% CI | p | N | β | 95% CI | p |
|  | 1 Year | 9686 | 0.07 | [0.05 to 0.09] | <0.001 | 9686 | 0.06 | [0.04 to 0.08] | <0.001 | 9672 | 0.08 | [0.06 to 0.1] | <0.001 | 7255 | 0.07 | [0.05 to 0.09] | <0.001 |
|  | 2 Year | 9363 | 0.07 | [0.05 to 0.1] | <0.001 | 9363 | 0.06 | [0.04 to 0.08] | <0.001 | 9348 | 0.07 | [0.05 to 0.09] | <0.001 | 7018 | 0.07 | [0.05 to 0.09] | <0.001 |
|  | 3 Year | 8694 | 0.06 | [0.04 to 0.08] | <0.001 | 8694 | 0.04 | [0.02 to 0.07] | <0.001 | 8681 | 0.06 | [0.04 to 0.08] | <0.001 | 6614 | 0.06 | [0.03 to 0.08] | <0.001 |
|  | 4 Year | 4047 | 0.02 | [-0.02 to 0.05] | 0.376 | 4047 | 0.01 | [-0.03 to 0.04] | 0.661 | 4041 | 0.02 | [-0.02 to 0.05] | 0.313 | 3174 | 0.02 | [-0.02 to 0.05] | 0.409 |

Abbreviations

N	Number of participants included in the analysis

ß	standardized effect estimate

95% CI	95% Confidence Interval

p	p-value

Results of the adjusted linear mixed models examining the association between baseline screen time and puberty timing are presented. Both the exposure (screen time) and the outcome (puberty timing) were standardized, and analyses were performed for males and females combined. Models were adjusted for fixed effects at baseline, including age, sex, race/ethnicity, highest parental education, total family income, Area Deprivation Index, BMI SDS, and weekly physical activity. Random intercepts were specified for study site and for families nested within sites.

Sensitivity analyses confirmed the robustness of the association between baseline screen time and puberty timing. Regardless of model specification, transformation of screen time (logarithmic vs. untransformed), or data source (youth-reported vs. parent-reported), and after excluding all participants with imputed covariates, a positive association between screen time and puberty timing was observed during follow-up years 1–3. In the fourth year, effects were smaller in all sensitivity analyses, and the 95% confidence intervals included zero.

| **Supplementary Table S12. Association Between Screen Time Group and Age at Mid-Puberty** | | | | | |
| --- | --- | --- | --- | --- | --- |
| **Screen Time Group (Screen Time Range, h/day)** | **Very Low (0.0 – 1.29)** | **Low (1.29 – 2.07)** | **Moderate (2.07 – 2.86)** | **High (2.86 – 4.0)** | **Very High (4.0 – 12.43)** |
| Age at mid-puberty, mean (SD) | 11.74 (1.41) | 11.66 (1.46) | 11.68 (1.47) | 11.56 (1.48) | 11.38 (1.46) |
| Age at mid-puberty – Difference Relative to Very Low Screentime Group in Months, mean (SD) | Ref. | -0.99 (17.49) | -0.74 (17.64) | -2.22 (17.76) | -4.35 (17.47) |
| Estimated Adjusted Effect of Screentime Group on Age Mid-Puberty, B in Months [95% CI] | Ref. | -1.22 [-2.07 to -0.38] | -0.84 [-1.70 to 0.02] | -1.76 [-2.64 to -0.89] | -2.47 [-3.38 to -1.56] |

Abbreviations

N	Number of participants included in the analysis

SD	Standard Deviation

Ref.	Reference Group

B	Effect Estimate; Estimated difference in age (years) at midpuberty compared with the reference group.

95% CI	95% Confidence Interval

Participants reaching mid‑puberty during follow‑up were stratified into five equal groups by baseline screen time. The table reports mean (SD), unadjusted differences relative to the Very Low group (in years and months), and adjusted differences as unstandardized coefficients (B) with 95% confidence intervals from linear mixed models. Models were adjusted for age, sex, race/ethnicity, parental socioeconomic information, physical activity, and BMI standard deviation score (BMI SDS), with random intercepts for study site and for families nested within sites. Negative B values indicate an adjusted mean difference in age at mid‑puberty (years) below zero relative to the Very Low screen time group

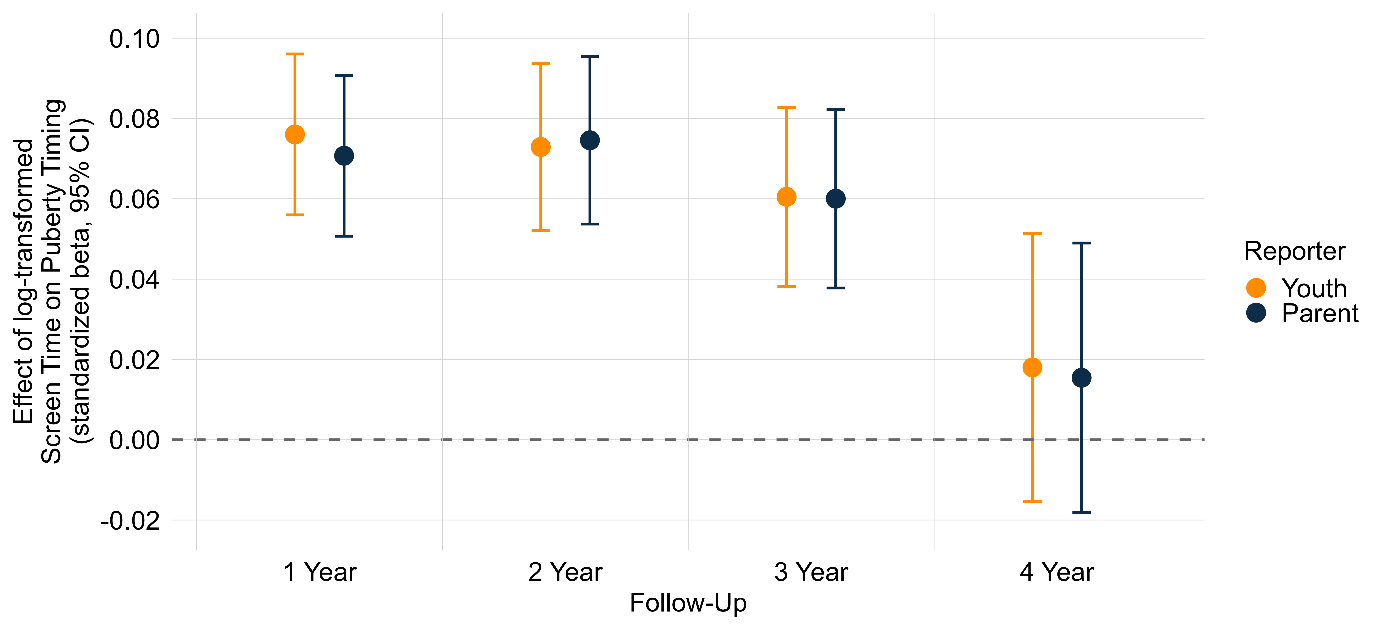

**Supplementary Figure S12. Sensitivity Analysis:** **Comparison of Estimated Effects of Parent- vs. Youth-Reported Screen Time** on Standardized Puberty Timing Across Follow-ups. Effect estimates are shown as points with 95% confidence intervals as lines. Yellow indicates results from adjusted linear mixed models using youth-reported, log-transformed screen time; blue indicates parent-reported screen time. Effect sizes were similar across both reporter types.

| **Supplementary Table S13. Further Analysis:** **Influence of Covariates on the Effect Estimate of Baseline Screen Time on Puberty Timing** | | | | | | | | | | | | | | | | |
| --- | --- | --- | --- | --- | --- | --- | --- | --- | --- | --- | --- | --- | --- | --- | --- | --- |
|  | **1 Year** | | | | **2 Year** | | | | **3 Year** | | | | **4 Year** | | | |
| **Adjustment Variation Model** | N | β | 95% CI | p | N | β | 95% CI | p | N | β | 95% CI | p | N | β | 95% CI | p |
| Crude | 9686 | 0.16 | [0.14 to 0.18] | <0.001 | 9363 | 0.15 | [0.13 to 0.17] | <0.001 | 8694 | 0.12 | [0.1 to 0.14] | <0.001 | 4047 | 0.04 | [0.01 to 0.08] | 0.008 |
| **Demografic Information (Demo)** | | | | | | | | | | | | | | | | |
| Sex | 9686 | 0.16 | [0.14 to 0.18] | <0.001 | 9363 | 0.15 | [0.13 to 0.17] | <0.001 | 8694 | 0.12 | [0.10 to 0.14] | <0.001 | 4047 | 0.04 | [0.01 to 0.07] | 0.009 |
| Sex, Age | 9686 | 0.16 | [0.14 to 0.18] | <0.001 | 9363 | 0.15 | [0.13 to 0.17] | <0.001 | 8694 | 0.12 | [0.09 to 0.14] | <0.001 | 4047 | 0.04 | [0.01 to 0.07] | 0.011 |
| Sex, Age, Ethnicity | 9686 | 0.12 | [0.10 to 0.14] | <0.001 | 9363 | 0.11 | [0.09 to 0.14] | <0.001 | 8694 | 0.09 | [0.07 to 0.12] | <0.001 | 4047 | 0.03 | [0 to 0.06] | 0.064 |
| **Parental Socioeconomic Information (SEI)** | | | | | | | | | | | | | | | | |
| Demo, Education | 9686 | 0.10 | [0.08 to 0.12] | <0.001 | 9363 | 0.10 | [0.08 to 0.12] | <0.001 | 8694 | 0.08 | [0.06 to 0.11] | <0.001 | 4047 | 0.03 | [0 to 0.07] | 0.044 |
| Demo, Education, Income | 9686 | 0.10 | [0.08 to 0.12] | <0.001 | 9363 | 0.10 | [0.08 to 0.12] | <0.001 | 8694 | 0.08 | [0.06 to 0.10] | <0.001 | 4047 | 0.04 | [0 to 0.07] | 0.043 |
| Demo, Education, Income, ADI | 9686 | 0.10 | [0.07 to 0.12] | <0.001 | 9363 | 0.10 | [0.08 to 0.12] | <0.001 | 8694 | 0.08 | [0.06 to 0.10] | <0.001 | 4047 | 0.03 | [0 to 0.07] | 0.051 |
| **Physical Factors** | | | | | | | | | | | | | | | | |
| Demo, SEI, Physical Activity | 9686 | 0.10 | [0.07 to 0.12] | <0.001 | 9363 | 0.10 | [0.08 t0 0.12] | <0.001 | 8694 | 0.08 | [0.06, 0.10] | <0.001 | 4047 | 0.03 | [0 to 0.07] | 0.053 |
| Demo, SEI, Physical Activity, BMI-SDS | 9686 | 0.07 | [0.05 to 0.09] | <0.001 | 9363 | 0.07 | [0.05 to 0.10] | <0.001 | 8694 | 0.06 | [0.04, 0.08] | <0.001 | 4047 | 0.02 | [-0.02 to 0.05] | 0.367 |
| Demo, SEI, Physical Activity, BMI-SDS. Puberty Timing | 9686 | 0.04 | [0.03 to 0.06] | <0.001 | 9363 | 0.06 | [0.04 to 0.08] | <0.001 | 8694 | 0.05 | [0.03, 0.07] | <0.001 | 4047 | 0.01 | [-0.02 to 0.04] | 0.527 |

Abbreviations

N	Number of participants included in the analysis

ß	standardized effect estimate

95% CI	95% Confidence Interval

p	p-value

The analysis of the contribution of individual covariates to puberty timing illustrates how the effect sizes of baseline screen time change with stepwise adjustment for demographic, socioeconomic, and physical factors. Linear mixed models examined the association between log-transformed baseline screen time and puberty timing at follow-up, with both variables standardized. Random intercepts were specified for study site and for families nested within sites, and analyses were performed for males and females combined. In the unadjusted (crude) model, effect sizes were positive and statistically significant during the first three years. With progressive adjustment, the effect sizes gradually decreased. Adjustment for ethnicity in particular led to a noticeable reduction in effects, whereas socioeconomic variables had only a minor influence on effect sizes. The most fully adjusted model, including all covariates, showed the smallest effects. Across the follow-up years, effects remained positive during years 1 to 3 but were markedly smaller in year 4, with 95% confidence intervals partially including zero.

| **Supplementary Table S14. Further Analyses: Sex-Specific Associations between Screen Time and Puberty Timing** | | | | | | | | | |
| --- | --- | --- | --- | --- | --- | --- | --- | --- | --- |
|  | | Male | | | | Female | | | |
| Follow-up | | N | ß | 95% CI | p | N | ß | 95% CI | p |
|  | 1 Year | 5068 | 0.05 | [0.02 to 0.07] | <0.001 | 4618 | 0.10 | [0.07 to 0.13] | <0.001 |
|  | 2 Year | 4930 | 0.06 | [0.03 to 0.09] | <0.001 | 4433 | 0.10 | [0.07 to 0.13] | <0.001 |
|  | 3 Year | 4624 | 0.05 | [0.02 to 0.08] | 0.003 | 4070 | 0.08 | [0.05 to 0.12] | <0.001 |
|  | 4 Year | 2159 | 0.00 | [-0.04 to 0.05] | 0.953 | 1888 | 0.03 | [-0.02 to 0.08] | 0.234 |

Abbreviations

N	Number of participants included in the analysis

ß	standardized effect estimate

95% CI	95% Confidence Interval

p	p-value

Results of the adjusted linear mixed models examining the association between baseline screen time and puberty timing are presented. Both the exposure (log-transformed screen time) and the outcome (puberty timing) were included as standardized variables. Models were adjusted for fixed effects at baseline, including age, race/ethnicity, highest parental education, total family income, Area Deprivation Index, weekly physical activity, and BMI-SDS. Random intercepts were specified for study site and for families nested within sites.

Among male participants, higher baseline screen time was consistently associated with later puberty timing during the first three follow-up years. In females, the effects were stronger over the same period. At the 4 year follow-up, effects for both sexes were smaller, and the 95% confidence intervals included zero.

| **Supplementary Table S15. Further Analyses: Sex-Specific Associations Between Screen Time Group and Age at Midpuberty** | | | | | | | | | | | |
| --- | --- | --- | --- | --- | --- | --- | --- | --- | --- | --- | --- |
|  | | **Males, N=6566** | | | | | **Females, N=4909** | | | | |
| **Screen Time Group (Screen Time Range, h/day)** | | **Very Low (0.0 – 1.29)** | **Low (1.29 – 2.07)** | **Moderate (2.07 – 2.86)** | **High (2.86 – 4.00)** | **Very High (4.00 – 12.43)** | **Very Low (0.0 – 1.29)** | **Low (1.29 – 2.07)** | **Moderate (2.07 – 2.86)** | **High (2.86 – 4.00)** | **Very High (4.00 – 12.43)** |
|  | Participants by Sex, N (%) | 570 (33.6) | 689 (40.6) | 713 (42.1) | 785 (46.3) | 809 (47.7) | 1125 (66.4) | 1006 (59.4) | 982 (57.9) | 910 (53.7) | 886 (52.3) |
|  | Age at Midpuberty in Years, mean (SD) | 12.85  (1.20) | 12.67  (1.25) | 12.73  (1.19) | 12.53  (1.27) | 12.30  (1.13) | 11.18  (1.15) | 10.96  (1.15) | 10.91  (1.14) | 10.72  (1.08) | 10.54  (1.00) |
|  | Age at Midpuberty – Difference Relative to Very Low Screentime Group in Months, mean (SD) | Ref. | -2.08  (15.03) | -1.35  (14.29) | -3.84  (15.22) | -6.56  (15.75) | Ref. | -2.62  (13.74) | -3.20  (13.71) | -5.55  (13.02) | -7.73  (11.97) |
|  | Estimated Effect of Screentime Group on Age Mid-Puberty, B [95% CI] in Months | Ref. | -0.84  [-2.31 to 0.62] | 0.04  [-1.44 to 1.53] | -0.91  [-2.38 to 0.56] | -1.46  [-2,97 to 0.06] | Ref. | -1.39  [-2.39 to -0.39] | -1.37  [-2.40 to -0.34] | -2.43  [-3.49 to -1.36] | -3.11  [-4.22 to -1,99] |

Abbreviations

N	Number of participants included in the analysis

SD	Standard Deviation

Ref.	Reference Group

B	Effect Estimate; Estimated difference in age (years) at midpuberty compared with the reference group.

95% CI	95% Confidence Interval

Participants reaching midpuberty during follow-up were stratified into five equal groups based on baseline screen time. The table shows means (SD), unadjusted differences relative to the Very Low group (in years and months), and adjusted differences as unstandardized coefficients (B) with 95% confidence intervals from linear mixed models, presented separately by sex. Models were adjusted for age, race/ethnicity, parental socioeconomic status, physical activity, and BMI standard deviation score (BMI SDS), with random intercepts for study site and families nested within sites. Negative B values indicate an adjusted mean difference in age at midpuberty below zero relative to the Very Low screen time group.

| **Supplementary Table S16. Further Analyses: Screen Time and Puberty Tempo** | | | | | | |
| --- | --- | --- | --- | --- | --- | --- |
|  | | N | Puberty Tempo, mean (SD) | ß | 95% CI | p |
|  | Overall | 9363 | 1.36 (1.33) | 0.04 | [0.02 to 0.06] | <0.001 |
|  | Male | 4930 | 0.91 (1.21) | 0.06 | [0.03 to 0.09] | <0.001 |
|  | Female | 4433 | 1.86 (1.27) | 0.04 | [0.00 to 0.07] | 0.026 |

Abbreviations

N	Number of participants included in the analysis

SD	Standard Deviation

ß	standardized effect estimate

95% CI	95% Confidence Interval

Results from adjusted linear mixed models examining the association between baseline screen time and puberty tempo over a 2-year follow-up. Both baseline log-transformed screen time and puberty tempo were standardized (z-scored) in the models. Puberty tempo was defined as the change in the Parental Pubertal Development Scale score over time (Δ PDS sum score score/year). In the overall sample (N=9363), higher baseline screen time was associated with a small increase in puberty tempo (ß=0.04 [95% CI, 0.02 to 0.06]). When stratified by sex, the effect remained modest in both males (N = 4930; ß=0.06 [95% CI, 0.03 to 0.09]) and females (N = 4433; ß = 0.04 [95% CI, 0.00 to 0.07]), indicating a generally small association between baseline screen time and puberty tempo for both sexes.

| **Supplementary Table S17. Further Analyses: Social Media Screen Time and Puberty Timing** | | | | | |
| --- | --- | --- | --- | --- | --- |
| Follow-up | | N | β | 95% CI | p |
|  | 1 Year | 9671 | 0.02 | [0.01 to 0.04] | 0.012 |
|  | 2 Year | 9345 | 0.00 | [-0.01 to 0.02] | 0.642 |
|  | 3 Year | 8678 | 0.01 | [-0.01 to 0.03] | 0.308 |
|  | 4 Year | 4040 | -0.01 | [-0.04 to 0.02] | 0.679 |

Abbreviations

N	Number of participants included in the analysis

ß	standardized effect estimate

95% CI	95% Confidence Interval

p	p-value

Adjusted standardized associations between baseline social media screen time and pubertal timing at follow-up years 1–4 from linear mixed models. The exposure (log-transformed social media screen time) and outcome (pubertal timing) were standardized. Models were adjusted for baseline age, sex, race/ethnicity, highest parental education, total family income, Area Deprivation Index, weekly physical activity, and BMI standard deviation score, with random intercepts for study site and for families nested within sites. Estimates were small in magnitude across all follow-up years; in follow-up years 2–4, the 95% confidence intervals included zero.

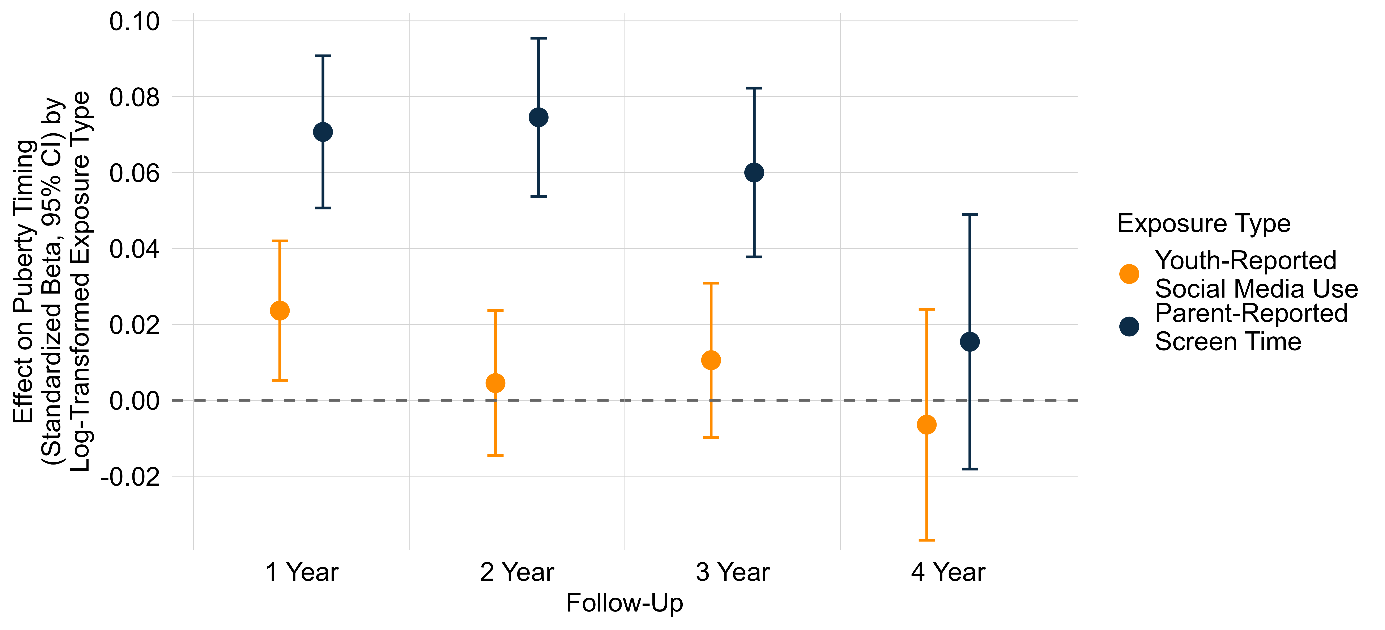
**Supplementary Figure S13. Additional Analysis: Comparison of Estimated Effects of Parent-Reported Screentime and Youth-Reported Social Media Use.** The standardized beta is shown as a point, with the 95% confidence intervals represented by lines. Results from the adjusted linear mixed models using youth-reported social media use are shown in yellow, compared with parent-reported screen time shown in blue.

*Software and packages*

All analyses were conducted in R (version 4.4.2). were fitted using the lme4 package (v1.1.36) [17] , with p-values calculated using the lmerTest package (v3.1.3) [18]. Multicollinearity was assessed by computing Generalized Variance Inflation Factors (GVIFs) using the car package (v3.1.3) [19].
